# Genome-wide DNA methylation patterns reveal clinically relevant predictive and prognostic subtypes in osteosarcoma

**DOI:** 10.1101/2020.11.26.20238584

**Authors:** Christopher E. Lietz, Erik T. Newman, Andrew D. Kelly, Santiago A. Lozano-Calderon, David H. Ebb, Kevin A. Raskin, Gregory M. Cote, Edwin Choy, G. Petur Nielsen, Benjamin Haibe-Kains, Martin J. Aryee, Dimitrios Spentzos

## Abstract

**Background:** Osteosarcoma (OSA) is an aggressive malignancy predominantly affecting children and young-adults. Genetic analysis has characterized very few recurrent mutations in OSA, and an improved understanding of interpatient tumor heterogeneity is needed for clinical management.

**Methods:** We analyzed genome-wide DNA methylation in primary OSA tumors from the NCI Therapeutically Applicable Research to Generate Effective Treatments (TARGET) program (n = 83) profiled using the Illumina 450K methylation array. We tested if broad genomic methylation predicted outcomes and defined supervised methylomic signatures predictive of Recurrence Free Survival (RFS), Chemotherapy Response (CR), and Metastatic disease at Diagnosis (MetDx). We assessed methylation pattern reproducibility in two independent clinical datasets (n = 28 and 34) and in an *in vitro* dataset (n = 11). Correlations between genomic methylation and transcription were tested using TARGET RNA-seq data. An *in silico* pharmacogenomic screen was performed to identify agents for future stratified application.

**Results:** Genome-wide methylation defined two subgroups. Relatively hypomethylated tumors experienced better chemotherapy response (Odds Ratio = 6.429, Fisher’s p = 0.007), longer RFS (metastatic, median 2.3 vs 26.7 months, localized, median 63.5 vs 104.7 months, stratified log-rank p = 0.006), and Overall Survival (p = 5×10-4) than hypermethylated tumors. Robust genomic methylation signatures predictive of RFS and CR were defined, and the signatures’ methylation patterns were reproducible in the independent clinical and *in vitro* datasets. The RFS signature was enriched for intragenic sites, whereas the CR signature and clinically relevant genome-wide methylation patterns were enriched for intergenic sites. Normal-tissue-like methylation patterns were associated with poor prognosis and *in vitro* analysis suggested that the methylation signatures are associated with tumor aggressiveness. Downstream transcriptional analysis revealed that genes annotated to the RFS methylation signature were also predictive survival. The transcriptional program represented in the RFS signature included several critical cellular pathways, whereas the CR signature was associated with much fewer known pathways, possibly reflecting a much broader cellular “methylation state” related to chemoresponse. A pharmacogenomic screen identified potential therapies, including epigenomic modifiers, for future stratified clinical application.

**Conclusion:** Genomic methylation offers insight into patient prognosis and could be a useful tool for developing alternate adjuvant therapeutic strategies.

## BACKGROUND

Osteosarcoma (OSA) is a rare primary bone tumor with a high metastatic potential with spread to the lungs even after treatment. The systemic management of pediatric OSA, which includes multi-agent chemotherapy, resulted in significant improvements in cure rates following adoption in the 1980s[1, 2], but has remained largely unchanged over the past 20 years; additionally, there is no proven second-line regimen for poor responders. The five-year survival rate remains at best 70% and it is much lower for patients who present with or progress to metastatic disease[3-7]. While recurrent germline and somatic mutations (including in *RB1, TP53*, and *ATRX*)[8-15] and unbalanced chromosomal rearrangements (including allele loss on 3q, 13q, 17p, and 18q)[16, 17], have been identified, there is no characteristic OSA translocation and significant genetic heterogeneity exists, posing a challenge with respect to diagnosis and identification of novel treatment targets. Furthermore, clinical assessment of chemotherapy response and identification of patients who would benefit from additional-line agents is currently reliant primarily on histologic tumor necrosis[6, 18], an imperfect surrogate which can only be assessed after multiple rounds of therapy have been administered[19-21]. Recently, when pathologically assessed response to chemotherapy was used as a marker to stratify patients for alternate or intensified adjuvant therapies in a large international clinical trial (EURAMOS), it did not result in a survival benefit[6, 22, 23]. This failure underscores the need for improved prognostic biomarkers to assist in the development of new therapies.

Recent work has focused on posttranslational modifications and epigenetic alterations as potential prognostic markers and therapeutic targets. Our group has demonstrated that microRNA (miRNA) expression patterns, including 14q32 non-coding locus-specific and genome-wide signatures, are predictive of clinical outcome and capture prognostic information distinct from that conveyed by pathologic necrosis and/or metastatic status[24-27]. We have also previously examined the modulation of miRNA expression by DNA methylation at 14q32[25, 27], but the significance and applicability of *global* methylation patterns in OSA remains much less clear.

Induction of global hypomethylation has been demonstrated to result in chromosomal instability and OSA formation in animal models[28], and *in vitro* treatment with demethylating agents has been shown to reverse epigenetically silenced tumor suppressor genes and inhibit OSA cell growth[29-31]. These observations are consistent with data showing that DNA methylation inhibitors preferentially affect genes that are expressed in normal tissue and silenced in cancer[32-34]. Genome-wide methylation signatures in clinical samples have been utilized to distinguish OSA from Ewing or synovial sarcoma, underscoring the notion of epigenetically-modified molecular phenotypes[35], and global methylation patterns have been shown to be prognostic of relapse risk in a small OSA clinical cohort[36].

In a rare and mostly pediatric tumor, like OSA, one limiting factor is the lack of easily accessible large and well annotated specimen cohorts. In this respect, we leveraged the large NCI TARGET dataset (Therapeutically Applicable Research to Generate Effective Treatments), which is a clinically-annotated, multi-omic dataset, recently released by the NCI in order to facilitate genomic research in OSA. We perform extensive bioinformatic analysis of the TARGET data and present analyses of genome-wide (and CpG site subtype-specific) methylation patterns identifying methylation signatures associated with clinical outcomes. We validate our findings in two additional independent datasets. Finally, making use of *in vitro* cell line and pharmacogenomic data, we identify candidate gene targets and drugs. In conclusion, we propose DNA methylation-based subtypes that warrant further exploration as a prognostic tool for patient risk stratification for novel therapies and for tailored therapeutic targeting and drug repurposing in OSA.

## METHODS

### Public data acquisition

We used the following publicly available previously published datasets (TARGET, Japanese National Cancer Center Research Institute (JNCCRI)[30], New York (NY)[35], and Genomics of Drug Sensitivity in Cancer (GDSC)[37-39])

All data analyzed was publicly available and experiments were approved by the presiding Institutional Review Board where each dataset was generated. All experiments were performed in accordance with relevant guidelines and regulations. Sample selection criteria are available in the source publications and at the TARGET initiative website (https://ocg.cancer.gov/programs/target/projects/OSA), see acknowledgements below. The TARGET dataset was downloaded from the TARGET data matrix (https://ocg.cancer.gov/programs/target/data-matrix). The JNCCRI (GSE125645) and NY dataset (GSE97529) were downloaded from the Gene Expression Omnibus.

### Illumina Infinium HumanMethylation450 BeadChip data processing

Each dataset assessed genome wide methylation with the Illumina Infinium HumanMethylation450 BeadChip platform. Array CpG annotations were obtained from the HumanMethylation450 v1.2 manifest file available at https://support.illumina.com/downloads/infinium_humanmethylation450_product_files.html. We downloaded pre-processed β values (methylated signal / methylated signal + unmethylated signal) for each clinical dataset. Technical details of β value generation are available in the source publications and at the TARGET initiative website (https://ocg.cancer.gov/programs/target/target-methods). Analysis of the TARGET dataset was performed using the 83 samples for which survival information was available. Data generated by probes containing frequent SNPs and those targeting the sex chromosomes were omitted from the preprocessed TARGET dataset[40], and we did not consider them in our analysis of the JNCCRI, NY, or GDSC datasets. Data was additionally processed by converting β values to M values and applying inter-array normalization via the *methylumi* R package[41]. M values were used for all analyses except where explicitly stated. M values have been shown to better meet assumptions for various parametric statistical tests than β values[42]. A filter selecting the 5% most variant CpG sites across the TARGET dataset was used for discovery analysis to reduce statistical noise. JNCCRI and NY methylation datasets were processed the same way. We downloaded raw signal intensity data (idat files) for OSA cell lines available in the GDSC dataset. Detailed methodology is available in the source publication. M values were then generated via the *minfi* R package[43] using Functional Normalization (an Illumina 450k array adapted quantile normalization) with default settings[44].

### Gene transcriptional analysis

RNA sequencing data was available for the TARGET dataset. Detailed methodology can be found on the TARGET initiative website (above). In summary, RNA was poly-A purified, gene libraries were prepared and multiplexed following standard Illumina protocol, and sequenced using the Illumina HiSeq 2000 platform. Transcript abundance was quantified with kallisto[45]. Count-level data was downloaded then normalized and log base 2 transformed via the DESeq2 R package[46].

Affymetrix Human Genome U219 transcription array data was available for the GDSC cell line dataset. Robust Multi-array Average (RMA) normalized transcription array data for the GDSC cell line dataset was downloaded from the project website (above)[47]. Detailed methodology can be found in the source publication.

The TARGET dataset assayed miRNA abundance using the ABI TaqMan Megaplex human miRNA qRT-PCR platform. Technical details are available on the TARGET initiative website (above). We transformed and normalized raw miRNA qRT-PCR data using standard 2^-ΔCt^ transformation as previously described[48]. Genes were considered potentially regulated by the methylation markers using the array CpG annotations provided in the Illumina HumanMethylation450 v1.2 manifest file (above). Illumina assigned CpG sites to genes based on the overlap of CpG and gene coordinates from the UCSC database[49] in the GRCh37 reference genome.

### Unsupervised and Supervised Survival analysis and prediction

All hierarchical clustering analysis was performed with the centered correlation and average linkage method and resulting subgroups were then analyzed for survival differences[50]. Cluster reproducibility was assessed with the R-index[51]. Heatmaps display mean centered and standard deviation scaled methylation patterns. Methylation based supervised analysis was performed using an average β approach whereby the average β value for each sample was calculated, and then samples classified into two groups based on the median average β value. Survival differences between cluster or average β defined groups were tested with Kaplan-Meier (KM) analysis and the log rank test for significance. MiRNA based supervised analysis was performed using the signed average approach as previously described[25, 27]. Multivariate analysis for confounding prognostic factors was performed using a Cox Regression model, with the methylation signature and relevant factors entered as independent co-variates.

### Differential methylation / transcription, gene set analysis, and other standard statistical tests

Two group continuous variable differentially analysis was performed by a t-test with p values adjusted to control the false discovery rate (FDR) using the Benjamini-Hochberg step-up procedure for multiple testing[52]. Gene set analysis for association with outcome was performed with the functional class scoring method, applying the LS/KS test with appropriate permutation-based p values[53]. Associations between two categorical variables were evaluated with two-tailed chi square / Fisher’s exact test and Odd Ratio (OR). Cramer’s V test was used to assess the strength of the classification concordance between different profiles. Pearson’s r statistic was used to evaluate continuous variable correlations. The hypergeometric test was used to test enrichment or depletion.

To assess specific associations between the drugs the methylation signatures and cell line aggressiveness and response to therapy *in vitro* we performed a simulation test using ten sets of 374 CpG sites (the number of sites in the outcome signatures) randomly selected from the sites found most variable in the clinical dataset. We then correlated methylation of the random CpG sites with cell line aggressiveness metrics and response to therapy.

### Methylation profile pathway enrichment analysis

Enrichment of Gene Ontology (GO) terms, BioCarta and KEGG (Kyoto Encyclopedia of Genes and Genomes) pathways in the outcome profiles was assessed using the DAVID Functional Annotation Tool[54]. GO terms and pathways with an FDR < 0.1 were considered enriched. Gene set analysis of transcriptional association of the methylation signature regulated genes and pathways with outcome was performed with the functional class scoring method. Pathways with a LS or KS permutation p < 0.05 and LS and KS permutation p < 0.1 were considered significant.

### Integrative pharmacogenomic analysis

We used the genes regulated by the RFS signature (CpG methylation-gene transcription |r| > 0.3, p < 0.001, N = 47) drug discovery using the PharmacoDB interface, which performs data analysis via the PharmacoGx R package[55, 56]. The 47 genes were entered in the PharmacoDB pipeline analyzing gene-drug predictive interactions from seven large scale datasets including a total of 650,894 individual drug sensitivity experiments, 1,691 cell lines, and 19,933 gene markers via a multivariate regression model adjusting for tissue source and experimental batch. We chose drug candidates at a stringent 0.001 regression p-value for predictive association with gene markers, and a regression coefficient > |0.15| (coefficient > |0.1| for hypothesis driven analysis) for effect size in a pan-cancer analysis, which used all cell lines in the database for increased sensitivity compared to the few OSA cell lines. To increase the specificity of the resulting drug list, only drugs passing the p-value and effect size filters for at least three genes regulated by the RFS signature were retained for further analysis. The *in vitro* experimental data of the filtered drug list was then evaluated specifically in the 11 OSA cell lines contained in the GDSC dataset[37] through the PharmacoDB Batch Query to obtain sensitivity measures (IC50 dose response metric) for response to candidate drugs. We assessed gene-drug correlations in the small subset of OSA cell lines and required at least two-thirds of the interactions to have the same correlation sign as those observed in the pan cancer (1,691 cell line) analysis. We finally selected drugs with the most variant IC50 response (top two tertiles) across OSA cell lines, and those more potent than cisplatin in at least two OSA cell lines.

To assess specific associations between the drugs the RFS methylation signature, we performed a random simulation test using the OS cell lines and ten sets of 374 CpG sites (the number of sites in the RFS signature) randomly selected from the 18,890 sites found most variable in the clinical dataset and not in the RFS signature. We then correlated methylation of the random CpG sites and drug response. Drugs were considered more associated with the methylation markers if the RFS signature contained more CpG sites strongly correlated with response (Pearson correlation, |r| > 0.6) than 80% of the sets of randomly generated CpG sites.

### Statistical software

The NCI BRB-ArrayTools v4.6.0, R (version 3.5.1)[57], R *ggplot2* package[58], R *minfi* package[43], R *methylumi* package[41], SPSS (version 24) software were used.

## RESULTS

### Genome-wide methylation patterns of primary OSA samples are associated with clinical outcomes

We identified three clinical datasets collectively composed of 132 samples analyzed by the same genome wide array-based methylation assay (Illumina Infinium HumanMethylation450 BeadChip). Clinical characteristics of the three cohorts, TARGET (n=83), Japanese National Cancer Center Research Institute[30] (JNCCRI, n = 34), and New York[35] (NY, n = 15), are presented in Table 1. 126 samples are biopsies or diagnostic resections collected prior to therapy. Six metastatic samples were also included in the JNCCRI dataset. The TARGET initiative sought to characterize pediatric cases, so it does not include older adult cases found in the JNCCRI and NY datasets. The JNCCRI and NY datasets included a larger fraction of axial cases.

**Table 1.** Clinical characteristic comparison of the TARGET, JNCCRI, and NY cohorts.

| Dataset | TARGET | JNCCRI | NY |
| --- | --- | --- | --- |
| N | 83 | 34<sup>1</sup> | 15 |
| Years collected | 2000 - 2012 | 1998 - 2013 | Not reported |
| Sex M / F | 47 / 36 | 16 / 15 | 7 / 8 |
| Age at diagnoses yrs. (min/median/max) | 3 / 14 / 32 | 7 / 13 / 68 | 6 / 15 / 80 |
| Tumor location (appendicular / axial) | 79 / 4 | 27 / 4 | 11 / 4 |
| Metastatic disease at diagnoses | 21 | 3 | Not reported |
| Pathologic response to chemotherapy<sup>2</sup><br>(optimal / suboptimal / unknown) | 20 / 22 / 41 | Not reported | Not reported |
| Chemotherapy treated | 83 | 30 | Not reported |
| MAP / AP | 12 |  |  |
| MAP and other | 5 |  |  |
| Not available | 66 |  |  |
| Events (OS Deaths / Recurrences) | 30 / 41 | 9 / 13 | Not reported |
| Follow up time<sup>3</sup> months<br>(min/median/max) | 12 / 56 / 144 | 4 / 69 / 240 | Not reported |

<sup>1</sup>28 primary OSA samples. Metastatic samples were analyzed for six patients.
<sup>2</sup>Optimal response defined as percent tumor necrosis $\geq 90\%$
<sup>3</sup>For censored patients

We first sought to discover if genome-wide methylation patterns offer insight into OSA clinical outcomes using the large, well annotated NCI TARGET dataset. Following standard pre-processing and normalization, we considered the 5% most variant CpG methylation sites (based on M values) in the dataset to remove statistical noise, leaving 19,264 sites for analysis. These most variant sites were enriched in the CpG Island Shore (Shore 4712 sites), Open Sea (7821 sites), and Enhancer (5231 sites) genomic regions (hypergeometric test p = 1.03×10^−3^, 7.31×10^−56^, and 1.33×10^−51^, respectively) and depleted in CpG Island (CGI, 5168 sites) and CpG Island Shelf (Shelf, 1563 sites) regions (p = 1.91×10^−53^, 1.38×10^−11^, respectively). It was interesting to observe that the depletion in CGI regions was largely due to a depletion of promoter associated CGIs (1230 sites, p = 1.19×10^−242^), and that sites in non-promoter associated CGIs (3938 sites) were enriched (p = 2.60×10^−19^) in the filtered dataset. Additionally, there was a large enrichment for CpG sites not annotated to any known gene (p = 1.03×10^−281^).

Unsupervised hierarchical clustering revealed two patient subgroups with strikingly different methylation patterns where one subgroup was largely hypermethylated relative to the other (Fig. 1A). The hypermethylated subgroup was enriched for tumors unresponsive to standard chemotherapy (pathologically assessed tumor necrosis in response to pre-operative chemotherapy (chemoresponse) < 90%, Odds ratio (OR) = 6.429, 95% CI = 1.662 – 24.860, Fisher’s Exact Test (2-sided) p = 0.007). The hypermethylated group displayed a trend for shorter Recurrence Free Survival (RFS) and Overall Survival (OS) in Kaplan-Meier analysis (Fig. 1B and E). However, when we stratified patients by the presence of metastases at diagnosis, the two main cluster groups were significantly associated with both RFS (p = 0.006) and OS (p = 5×10^−4^, Fig. 1C and F). The two main cluster groups were not different with respect to age (p = 0.531), metastasis at the time of diagnoses (MetDx, a known strong prognostic factor, p = 0.615), or sex (p = 0.661). We tested differential methylation between the two groups, found many strong associations (7075 sites, p < 0.001, Benjamini-Hochberg False Discovery Rate (FDR)[52] < 0.05), 77.0% of which had an average β value difference > 0.2. 98.9% of these sites were hypermethylated in the suboptimal chemoresponse cluster subgroup (Fig. 1A red). Large average β value differences (> 0.4) were found at 1.1% of sites, all of which were hypermethylated in suboptimal chemoresponse cluster subgroup. We found a strong significant enrichment for Shelf, Open Sea, and non-genic sites (p = 1.75×10^−59^, 2.40×10^−237^, 8.30×10^−30^, respectively). Sites on chromosomes 1, 7, 8, 13, 14, 15, and 22 were significantly (p < 0.05) enriched. Most striking was the prevalence of sites located in the 14q32 locus among the top sites. Nineteen of the 100 most differentially methylated sites were located within 14q32, including the most differentially methylated site, cg08175935 which is annotated to MIR127, MIR433, and RTL1. 14q32 is an imprinted region encoding many microRNAs[59, 60], the transcription and methylation status of which we have previously reported to be prognostic of patient survival[24, 25, 27].

**Figure 1.**
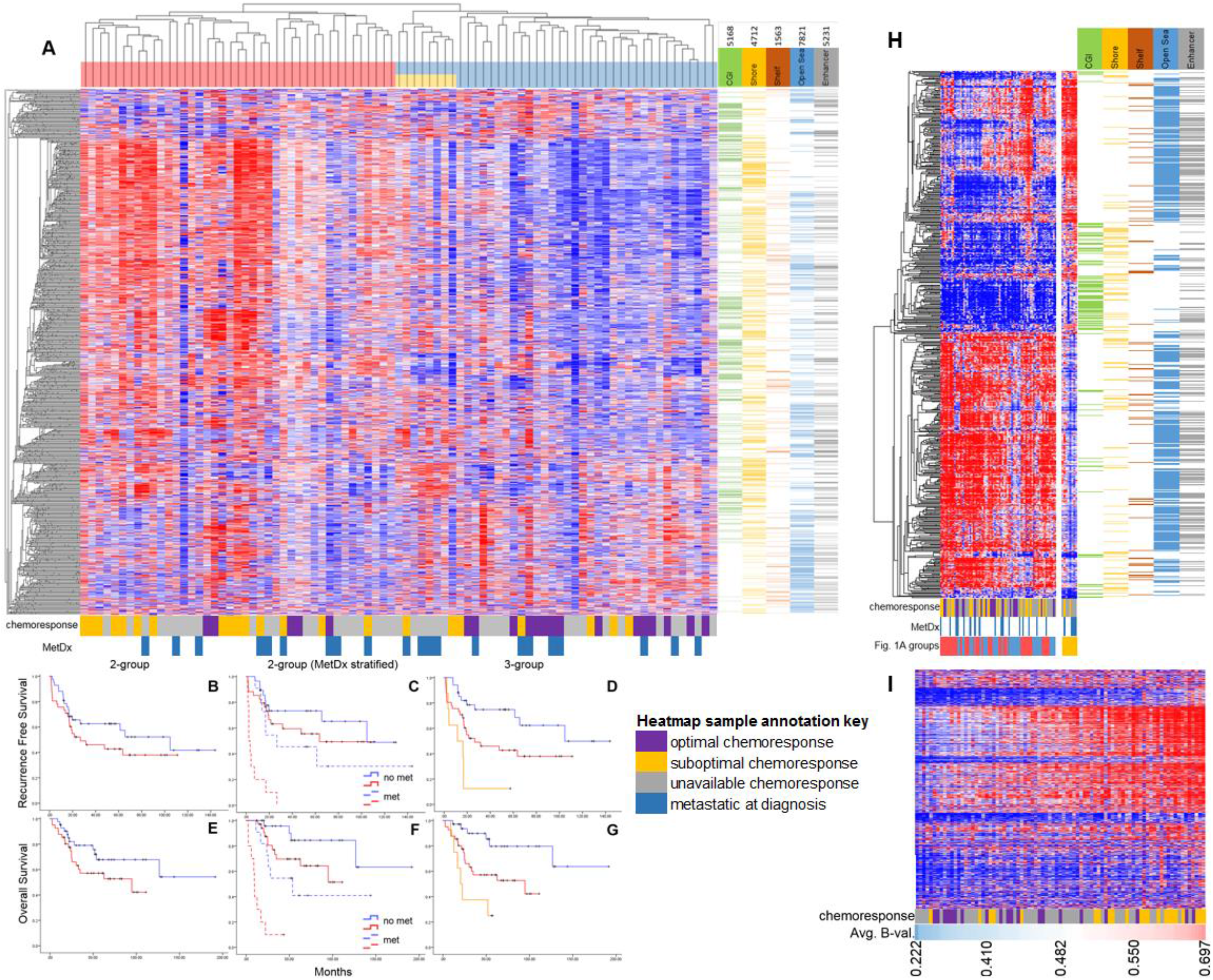
Global methylation map of primary tumors. **A)** Unsupervised hierarchical clustering of the TARGET samples using the 95% most variant CpG methylation sites. Annotations for CpG islands (CGIs) (green), Shore (yellow), Shelf (orange), Sea (blue), and Enhancer (grey) genomic regions are shown to the right of the heatmap. Cluster reproducibility (R) indices were high, 0.98 / 0.95 for 2 and 3 groups respectively. **B)** RFS analysis of the two primary cluster groups (median RFS = 26.7 and 104.7 mo., Log-Rank p = 0.176). **C)** MetDx stratified RFS analysis of the two primary cluster groups (not metastatic: median RFS = 63.5 and 104.7 mo., metastatic: median RFS = 2.3 and 26.7 mo., pooled Log-Rank p = 0.006). **D)** RFS analysis of the three primary cluster groups (median RFS = 11.5, 26.7, and 104.7 mo., Log-Rank p = 5.0 x 10^−4^). Pairwise RFS analysis of the three primary cluster groups. Short (11.5 mo.) vs. intermediate (26.7 mo.) median RFS Log-Rank p = 0.034. Intermediate vs. long (104.7 mo.) median RFS Log-Rank p = 0.022. Short vs. long median RFS Log-Rank p = 2.7 x 10^−5^. **E)** OS analysis of the two primary cluster groups (median OS = 94.7 mo. and Not Yet Reached (NYR), Log-Rank p = 0.106. **F)** MetDx stratified OS analysis of the two primary cluster groups (not metastatic: median OS = NYR and NYR, metastatic: median OS = 20.6 and 111.9 mo., pooled Log-Rank p = 5×10^−4^). **G)** OS analysis of the three primary cluster groups (median OS = 20.4, 94,7 mo., and NYR, Log-Rank p = 7.4 x 10^−4^. Pairwise OS analysis of the three primary cluster groups. Short (20.4 mo.) vs. intermediate (94.7 mo.) median OS Log-Rank p = 0.067. Intermediate vs. long (NYR) median OS Log-Rank p = 0.008. Short vs. long median OS Log-Rank p = 1.8 x 10^−4^. **H)** Supervised β value heatmap of the 987 CpG sites differentially methylated (FDR < 0.05) between the poor prognosis (yellow) and other cluster groups (red and blue) in **A. I)** Supervised heatmap of the 95% most genome-wide variant sites. Samples are ordered from low to high average β value. Two sample groups generated by a median split of the average β values were significantly associated with pathologic response to chemotherapy (Fisher’s exact p val. = 0.002, OR = 10.2 (95% CI: 2.5 - 41.7).

When we considered three, instead of two, main clustering groups, we noticed an increased RFS and OS discrimination (Fig. 1D and G). These three groups remained significantly associated with both outcomes when stratified for metastasis at the time of diagnosis. Although the third subgroup was a subset of the hypomethylated subgroup, its members had poor long-term survival. The only two samples in this subgroup with chemotherapy information had suboptimal response, and half of the patients in this subgroup presented with metastatic disease. We compared the methylation profiles of the small cluster group with exceptionally poor prognosis to the other samples and identified 972 differentially methylated sites (FDR < 0.05), 96.4% of which had an average β difference > 0.2, and 48.5% which were hypermethylated in the poor prognosis group. 14.8% of the sites had an average β difference > 0.4, of which 47.2% were hypermethylated in the poor prognosis subgroup. The differentially methylated sites were strongly enriched Open Sea and Enhancer regions (hypergeometric test p = 3.2×10^−39^, 2.3×10^−33^, respectively). Sites on chromosomes 1, 3, 8, 12, 13, and 14 were also significantly enriched (p < 0.05). Inspection of the β values at these sites revealed the poor prognosis group is hypermethylated a small number of CGIs and a subset of Open Sea sites, and hypomethylated at a larger subset of Open Sea sites (Fig. 1H).

We also sought to explore individualized patient outcome prediction using the global methylation patterns by applying a supervised average methylation approach. To do this, we simply averaged all β values for each sample and then ranked patients by this average methylation metric, creating two, three, and four patient groups using the median, tertile, and quartile average methylation values. We noticed these patient groups were significantly associated with response to chemotherapy (χ^2^ test, p = 0.002, 0.002, and 0.017 resp., Fig. 1I). We observed an inverse relation between global methylation and chemoresponse where hypomethylated samples were more likely to have optimal response and hypermethylated samples were more likely to have suboptimal response. Furthermore, we found evidence of a methylation stratified effect between methylation and chemoresponse when we performed pairwise comparisons between each of the groups, as all the odds ratios displayed a consistent and expected relationship (OR < 1). Only a trend for association between these same β value-based risk groups and RFS was observed (2-groups, KM log-rank p = 0.157), however, the groups were significantly associated with RFS when stratified for metastatic disease at diagnosis (KM log-rank p = 0.044).

We then investigated if methylation patterns of specific genomic regions differ from those observed at the genome-wide scale and if they offer additional insights into clinical outcome. We thus performed hierarchical clustering using CGI, Shore, Shelf, Open Sea, and Enhancer regions. The two main cluster groups generated using methylation of each of these regions were not associated with patient age (Fisher’s exact test, p > 0.05). CGI defined cluster groups exhibited a balance of relative hyper and hypo methylation (Fig. 2A), a stark contrast to the patterns observed across the entire genome (Fig. 1A), and patterns observed in CpG sparse regions (Open Sea, Fig. 2E). The methylation patterns of each region (except for Shelf regions) generated two groups of samples with very different response to chemotherapy (Fisher’s exact test, OR > 8, p < 0.005 for all three analyses). The suboptimal response subgroup was largely hypermethylated in the Shore, Open Sea, and Enhancer regions. Sample risk subgroups generated by CGI and Enhancer methylation patterns provided the strongest discrimination for RFS (Fig. 2B-D, L and M). CGI and Enhancer methylation sample cluster groups were also significantly associated with OS (Fig. 2E-G, N and O). The high and low risk subgroups defined by CGI patterns had a balance of hyper and hypo methylation. Hypomethylation of Enhancer regions was associated with longer RFS. When we stratified for metastases at the time of diagnosis, we observed that the cluster groups generated with each of the genomic regions were significantly (KM log-rank p < 0.05) associated with RFS and OS (Fig. S1). Average β value-based risk groups generated using only CGI or Enhancer sites were not associated with survival, unlike groups generated by clustering, given the mix of hyper and hypo methylation in most samples observed in Fig. 2. However, average β value-based risk groups generated with each region were significantly associated with chemotherapy response (p < 0.05) and the hypermethylated groups contained fewer responders.

**Figure 2.**
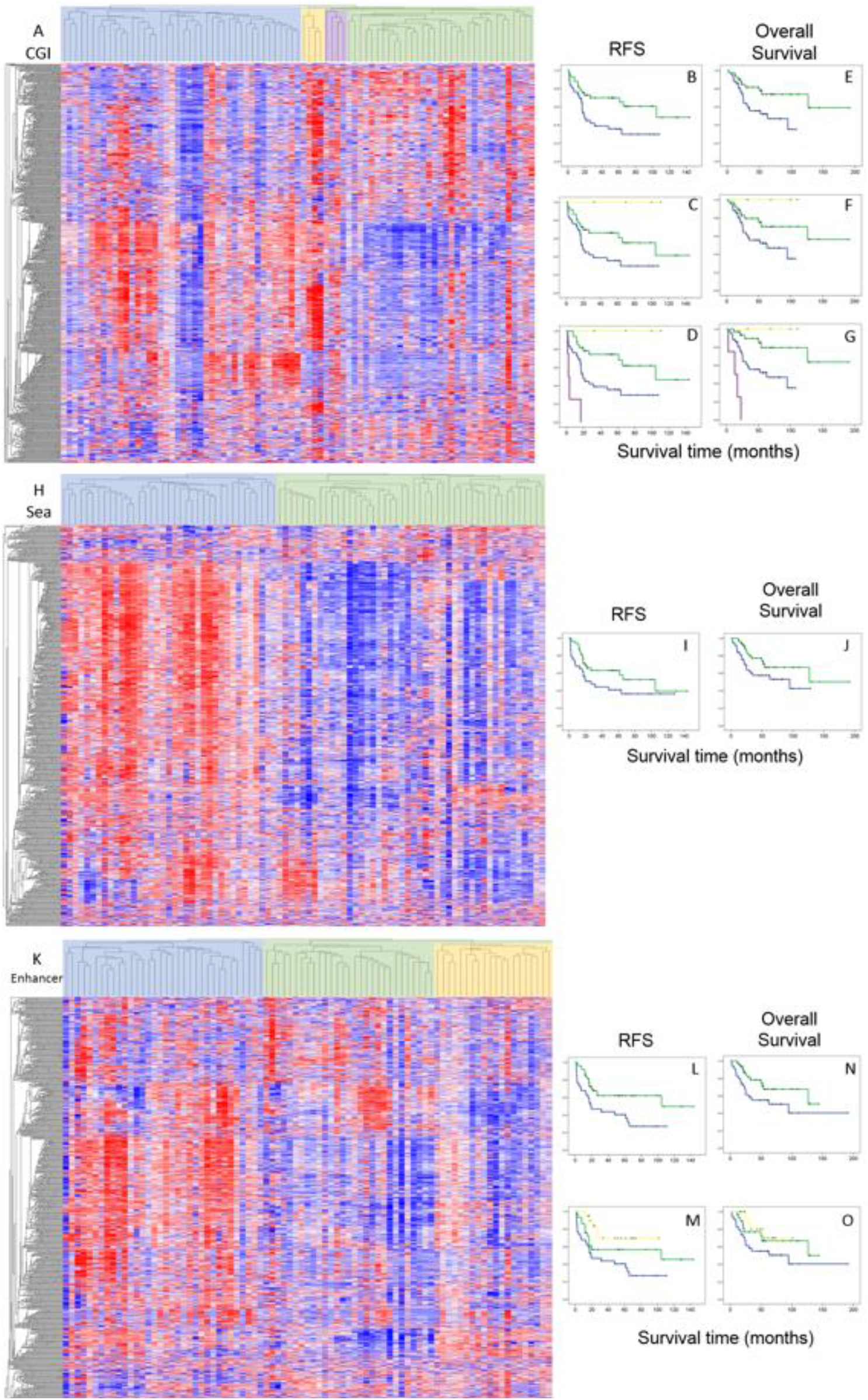
Methylation patterns of genomic regions. Clustering sample groups are displayed below the heatmaps. **A)** Unsupervised hierarchical cluster analysis using CGI methylation (R indices = 0.83, 0.92, and 0.80 for 2, 3, and 4 groups, respectively). **B-D)** RFS analysis of the 2, 3, and 4 primary CGI clusters (Log-Rank p = 0.009, 0.015, and 6.7 x 10^−7^, respectively) **E-G)** OS analysis of the 2, 3, and 4 primary CGI clusters (Log-Rank p = 0.016, 0.036, and 3.2 x 10^−8^, respectively). **H)** Cluster analysis using Open Sea methylation (R index = 0.72 for 2 groups). **I)** RFS analysis of the 2 primary Open Sea clusters (Log-Rank p = 0.074). **J)** OS analysis of the 2 primary Open Sea clusters (Log-Rank p = 0.084). **K)** Cluster analysis using Enhancer methylation (R index = 0.80 / 0.76 for 2 and 3 groups respectively). **L** and **M)** RFS analysis of the 2 and 3 primary Enhancer clusters (Log-Rank p = 0.008, and 0.015, respectively). **N** and **O)** OS analysis of the 2 and 3 primary Enhancer clusters (Log-Rank p = 0.045, and 0.116, respectively).

### Methylation signatures are associated with clinical outcomes in univariate probe level analysis

After observing that broad methylation patterns are associated with outcome, we sought to discover subsets of individual CpG methylation sites most strongly associated with outcome and thus potentially clinically applicable for prognostication. We analyzed individual probes for association with RFS and CR using both a univariate p value of 0.05 and a Benjamini-Hochberg FDR corrected p value of 0.1 as cutoffs to identify CpG methylation sites associated with the RFS and CR outcomes. There were fewer strong associations with MetDx, so we used a more relaxed cutoff (univariate uncorrected p < 0.05) to identify sites more tentatively associated with this outcome. With this approach, we found 885 CpG sites were associated with RFS (univariate p < 0.004), 6224 associated with CR (univariate p < 0.032) and 671 with MetDx. We observed a significant degree of overlap between the RFS and CR associated lists (hypergeometric test p < 0.001), so we proceeded to refine more specific outcome signatures by removing probes shared by more than one of these lists. This generated a list of 374 CpG associated with RFS, 5641 with CR, and 671 with MetDx (Table S1). Both the RFS and CR signature were significantly (hypergeometric test p < 0.05) enriched for sites in CpG sparse regions (Open Sea) and depleted for sites in CpG dense regions (CGI). The depletion of CGI associated sites was strongest for non-promoter CGIs in the RFS signature, and promoter associated CGIs in the CR signature. Additionally, the RFS signature was significantly enriched for sites in genic sequences, whereas the CR signature was significantly enriched for sites in non-genic sequences. The MetDx signature was significantly enriched for sites in genic sequences (complete results in Table S2).

We secondarily performed a global analysis for methylation associations with the more distant OS endpoint and identified 149 sites significantly associated (p < 0.05, FDR < 0.1) with this outcome, 93.3 and 29.5% of which were members of the 885 and 374 RFS associated site lists, respectively. In a targeted manner, we tested the RFS associated sites for association with OS. Not surprisingly, we found 97.6 and 95.7 % of the 885 and 374 RFS sites were also significantly associated (p < 0.05, FDR < 0.1) with OS.

Then using the refined outcome signatures, we performed semi-supervised hierarchical clustering to visualize the methylation patterns across the dataset (Fig. 3). The two primary patient risk subgroups were not different with respect to age or sex (Fishers’s exact test p > 0.05) when each of the signatures was used for hierarchical clustering analysis. Patient risk subgroups generated using the RFS and MetDx signatures displayed a balance of hypo and hypermethylation. This pattern was especially pronounced with the RFS signature, where the poor prognosis subgroup was defined by predominantly hypomethylated CGI’s and hypermethylated Open Sea genomic regions. It was interesting to observe that the RFS signature predicted a few patients with sub-optimal chemotherapy response to achieve long-term survival as it is known that a small subset of patients who do not respond to therapy can still have good outcomes. The CR signature displayed similar methylation patterns to those seen in the genome-wide analysis (Fig. 1A), with a largely hypermethylated poor chemoresponse group and a hypomethylated optimal chemoresponse group. Notably, of the small subset (13) of CR signature CpG sites with an opposite methylation-chemoresponse association, 11 were in CpG islands, and the other two were in neighboring Shore regions.

**Figure 3.**
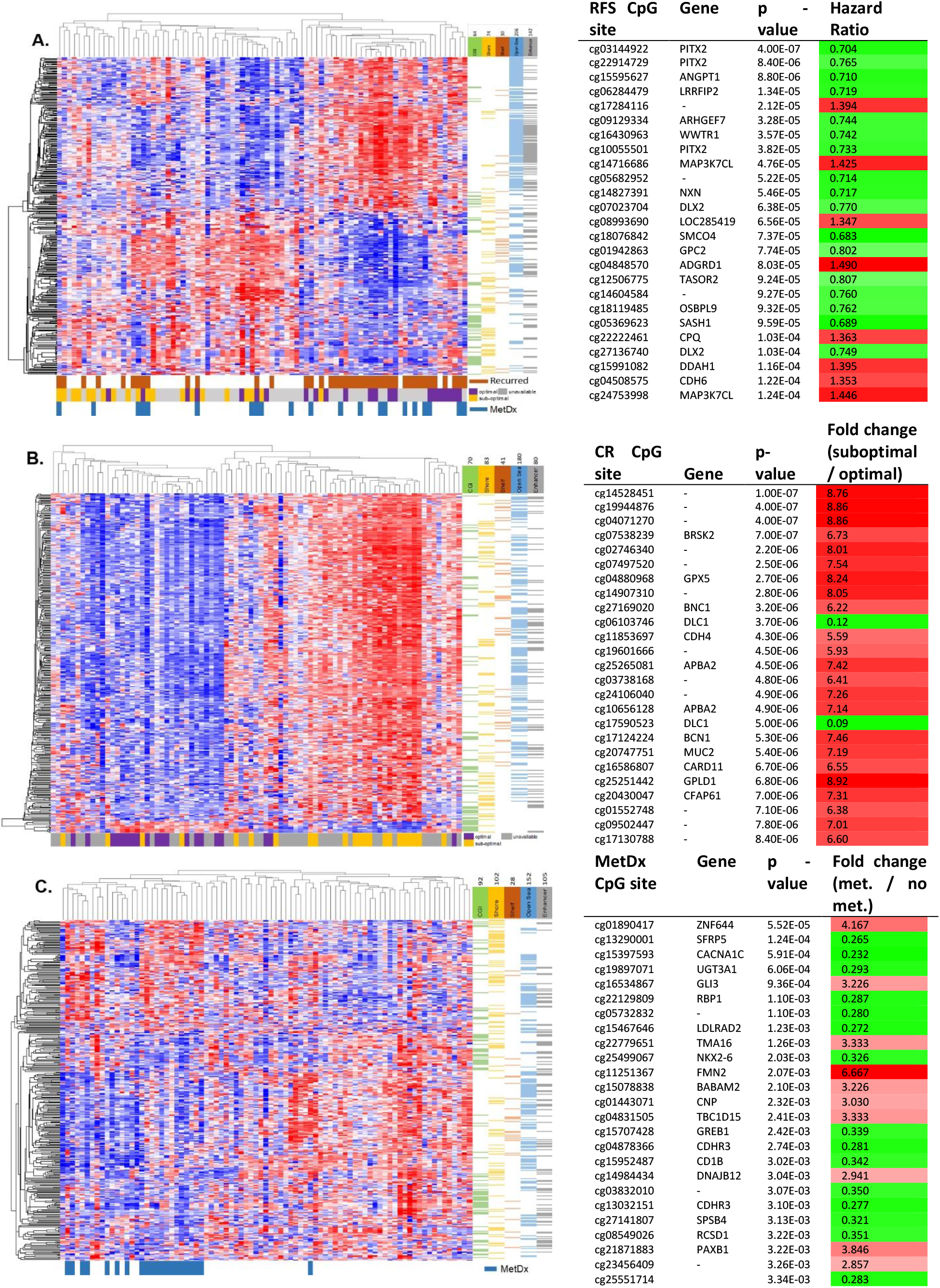
Supervised methylation signatures associated with outcome. Left panel) Semi-supervised clustering analysis using the RFS **(A)**, CR **(B)**, and MetDx **(C)** outcome signatures (2 group cluster R-indices = 0.846, 0.862, and 0.848 respectively). Right panel) The 25 CpG methylation sites in each signature most significantly associated with the respective outcome in univariate analysis are shown in the figure. RFS **(A)** and CR **(B)** associations were significant at the FDR < 0.05 level, whereas MetDx **(C)** associations were significant only at the univariate level.

We were also interested to see if the methylation patterns of the outcome signatures produced similar sample classifications to those generated with global patterns. The CR signature generated sample classifications highly concordant with the global methylation patterns (Cramer’s V = 0.783, Fisher’s exact test p = 1.18×10^−13^). The RFS and MetDx signatures also classified samples similarly to the global methylation patterns (RFS v. Global: Cramer’s V = 0.255, Fisher’s exact test p = 0.026, MetDx v. Global: Cramer’s V = 0.476, Fisher’s exact test p = 1.90 x10^−5^), although the associations were not as strong, indicating long term outcome and tumor aggressiveness may be mediated by different and more specific epigenetic mechanisms than response to chemotherapy.

While we avoided testing potential associations between the outcome defined methylation signatures and their respective outcome out of concern for overfitting, to illustrate the effect size of the constituent CpG sites in the RFS signature, we performed KM analysis using patient groups defined by methylation value quantiles of select sites of possible biologic interest ranked close to the top of list (Fig. S2). We observed a potential methylation level stratified relationship between methylation of these sites (and others) with RFS, whereby more extreme levels of methylation were associated with more extreme survival times.

### Methylation patterns are predictive of RFS independent of known prognostic factors

Pathologic necrosis following pre-operative chemotherapy is the single validated prognostic factor in OSA. Given the observation that there was discordance between chemoresponse and the patient risk groups classified by the RFS signature, we sought to test if the methylation markers predicted survival independent of chemoresponse. We observed that 30.7% and 23.5% of the RFS signature and all CpG sites found associated with RFS, respectively remained significantly predictive of RFS over and above chemoresponse in a multivariate Cox regression, despite the analysis being performed on the smaller subset of samples (n = 42) with chemoresponse information (Table S1). We analyzed the over/under representation of genomic regions in the subset of the RFS signature predictive of RFS independent of chemoresponse and found that CGI’s were enriched (hypergeometric test, p = 6.71×10^−7^) and Open Sea regions were depleted (p = 3.83×10^−6^) relative to the complete signature.

Our group, and others, have also proposed that miRNA transcriptional profiles could be a useful prognostic marker for this disease[24, 25, 27]. Therefore, we used the microRNA assays that were reported for the same specimens in the TARGET dataset to perform another multivariate analysis which demonstrated 100% of both the RFS signature and all CpG sites found associated with RFS were predictive of RFS over and above chemoresponse (Table S1).

### Methylation patterns are reproducible in two independent datasets

We next examined two smaller independent publicly available datasets (JNCCRI[30] and NY[35]) also providing Illumina 450k array data to test if we could reproduce the methylation patterns discovered in the TARGET dataset. Because chemoresponse and survival data were not available in these public cohorts we could not directly test the prognostic and predictive value of our signatures. We instead investigated if the pre-defined methylation signatures could classify samples similarly and displayed similar methylation patterns to those observed in the TARGET dataset. To do this, we first observed the distribution of the signatures’ average β values across samples (Fig. 4A) and found them to be similar by broad inspection, albeit with a trend towards hypermethylation in the NY dataset. We also performed semi-supervised hierarchical clustering using the TARGET defined global (unsupervised) and outcome (supervised) signatures in the independent datasets and analyzed the methylation patterns of the two primary cluster groups in each analysis (Fig. 4B-G). In both independent datasets, the RFS and MetDx signatures displayed hypo and hypermethylation in each risk group, and the CR signature was almost entirely hypermethylated in one group relative to the other, exactly as was observed in the TARGET data. Sample subgroups generated with each of the methylation signatures in each dataset were not different with respect to age (Fisher’s exact test p > 0.05), except for those generated using the RFS signature in the JNCCRI dataset (p = 0.03).

**Figure 4.**
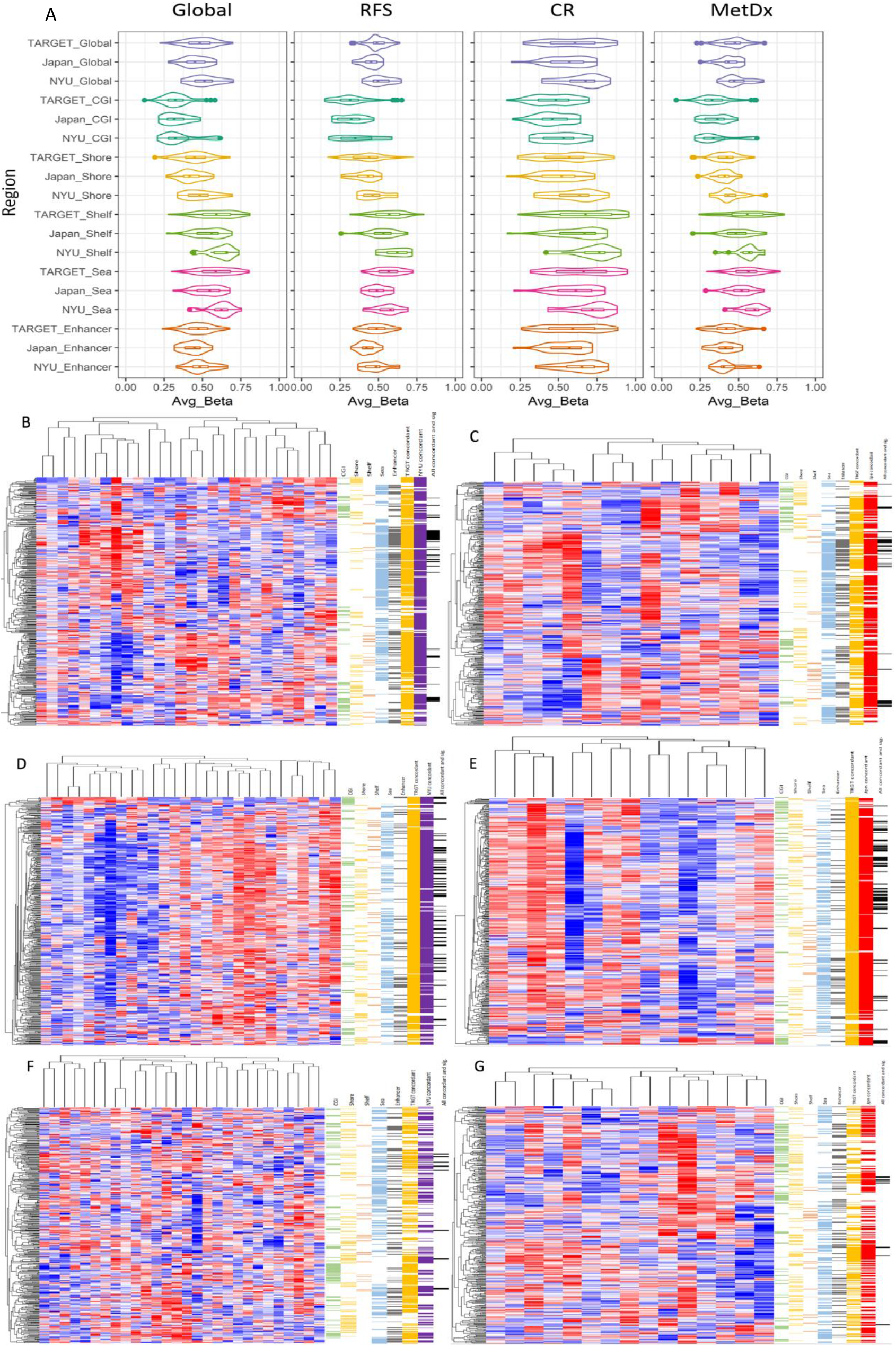
Methylation patterns of the outcome signatures in two independent datasets. **A)** Average β value distributions of global methylation and the outcome signatures. **B-G)** Hierarchical clustering of the RFS (top panel), CR (middle panel), and MetDx (bottom panel) signatures in the JNCCRI (left) and NY (right) independent datasets. Genomic regions (first five color bars: CGI-green, Shore-yellow, Shelf-orange, Open Sea-blue, and Enhancer-grey) are shown to the right of each heatmap. CpG sites displaying concordant hypo / hyper methylation patterns in each of the independent clinical datasets (last two color bars: TARGET-orange, JNCCRI-red, NY-purple) are also annotated to the right of each heatmap. Detailed concordance results are presented in **Table S3**.

We performed differential methylation analysis between the two primary cluster subgroups in each dataset and calculated the fraction of CpG sites differentially methylated (p < 0.05) between cluster groups which had concordant fold change directions in both the discovery and validation datasets (Fig. 4 and Table S3). We observed that the methylation patterns in the independent JNCCRI and NY datasets were remarkably similar to those observed in the TARGET dataset. This was especially true for the outcome signatures.

The JNCCRI dataset contained unique information for a few normal bone (2), lung (1), and metastatic (6) samples, which, although of limited sample size, offered some additional insights. We observed that the average methylation level of CGIs was elevated, and the average methylation level of Open Sea regions was depressed in tumor compared to normal tissue, a relationship commonly observed in other cancers (Fig. 5A)[61]. Furthermore, the methylation patterns indicated that the outcome signature predicted that tumors with more normal-like methylation patterns to be more aggressive/less responsive to therapy. We also found Shore patterns mirrored those of CGIs, and Shelf patterns mirrored those of the Open Sea regions. While most CGI’s are hypomethylated in normal tissue[62], the CGIs in the CR signature were found to be relatively hypermethylated in normal tissue. Furthermore, the CR signature in general was relatively hypermethylated in normal tissue, suggesting that tumors with more normal-like methylation patterns are less likely to respond to therapy.

**Figure 5.**
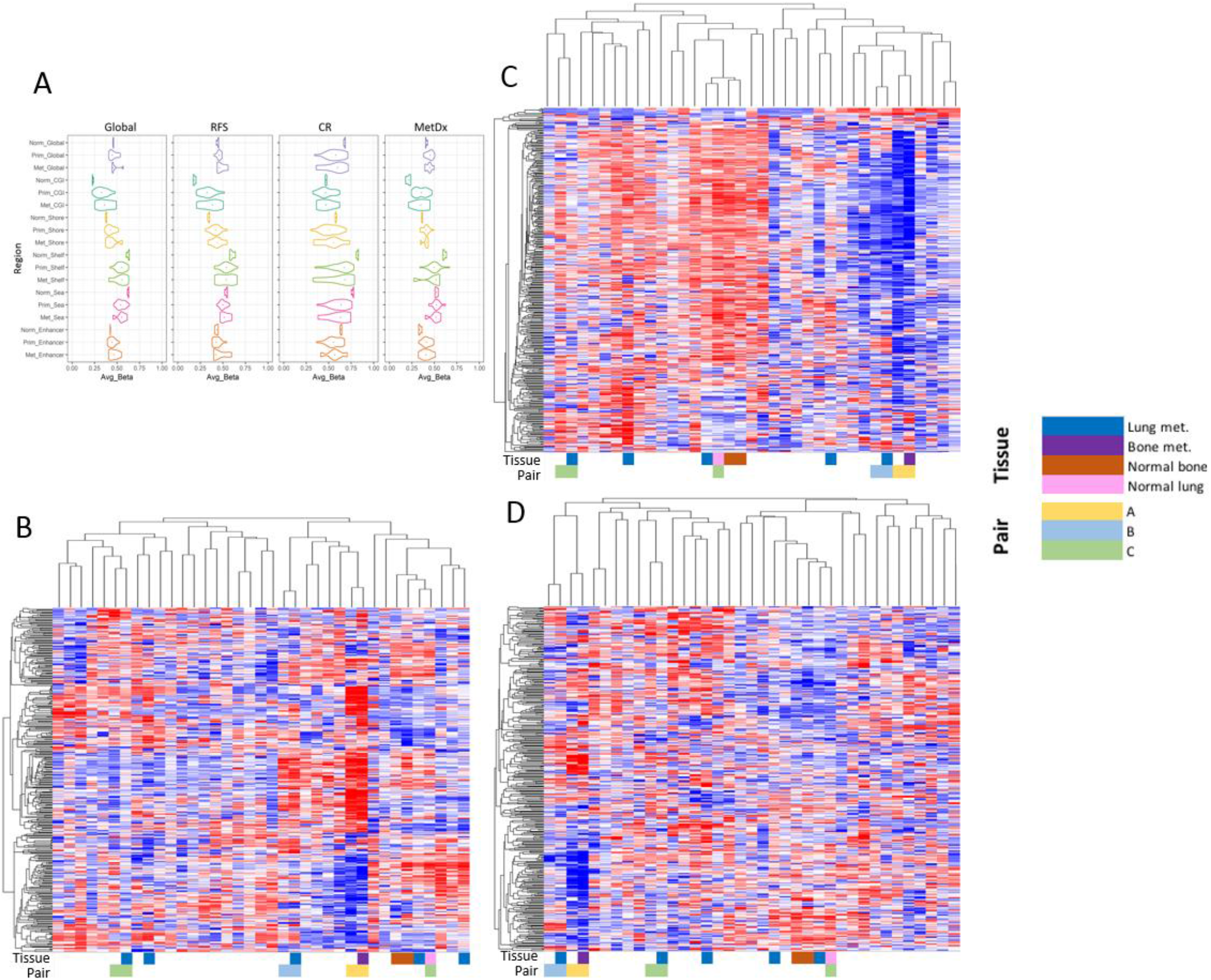
Methylation map including primary as well as metastatic tumor and normal tissue. **A)** Plots displaying the average β value distributions of global methylation, outcome signatures, and genomic region subsets for normal, primary, and metastatic samples in the JNCCRI dataset. **B-D)** Semi-supervised hierarchical clustering of normal, primary, and metastatic samples using the RFS, CR, and MetDx signatures, respectively. Sample tissue type and paired samples are displayed underneath the heatmaps.

Additionally, while the RFS and MetDx signature were largely hypermethylated in tumors, the CR signature was hypomethylated in tumors, indicating these methylation signatures may be tracking distinct elements of biology. We also performed semi-supervised hierarchical clustering using the outcome signatures with normal, primary, and metastatic samples (Fig. 5B-D). Normal tissue samples always clustered together. Metastatic samples did not cluster together, however primary-metastatic tissue pairs were always grouped as more similar to each other than any other sample. This raises the possibility that the specific outcome signatures may not substantially change with metastatic tumor progression, though this should be validated in future larger dedicated cohorts.

### Methylation offers insight into potential targeted application of immune checkpoint inhibitors

CpG sites from a previously reported epigenetic signature (EPIMMUNE)[63] generated using another Illumina methylation platform (EPIC array) for response to the immune checkpoint inhibitors nivolumab and pembrolizumab in non-small cell lung cancer were enriched in the TARGET dataset’s most variant sites (hypergeometric test, p = 1.94×10^−14^). The EPIMMUNE signature contains 301 CpG sites, 128 of which were interrogated by the lower resolution 450k array and passed the pre-processing criteria in the TARGET dataset. These sites were not enriched in the outcome signatures, but we did find through differential methylation analysis that 27 of them were univariately associated with response to chemotherapy (FDR < 0.05). Additionally, hierarchical clustering of the TARGET dataset using the EPIMMUNE signature revealed that a subset of OSA samples had methylation patterns similar to the lung cancer samples that responded to immunotherapy (Fig. S3). Notably, these samples belong to the hypermethylated, poor chemoresponse group (Fig. 1A, red).

### Biologic and functional annotation of the methylation profiles

In order to gain insight into the methylation profile content we used the DAVID Functional Annotation Analysis tool (Table S4)[54]. We first tested enrichment in CGI and Enhancer regions. The seven most enriched Gene Ontology (GO) terms enriched in the CGI profile related to transcription, e.g. “positive regulation of transcription from RNA polymerase II promoter” (Table S4). Additional enriched terms, such as “embryonic skeletal system morphogenesis”, and other developmental pathways, were also identified. The Enhancer regions were also enriched with terms relating to transcription and development, as well as cell adhesion. Pathways enriched in the RFS signature included processes such as “positive regulation of endothelial cell migration”, “positive regulation of activated T cell proliferation”, “BCR signaling”, and multiple pathways pertaining to neuronal activity. The RFS signature was also enriched for CpG sites on the chromosome “1q22-23” and chromosome “6p21-22” cytogenetic bands. Notably, these are the two largest tRNA clusters in the human genome (encoding > 50% of tRNAs)[64], and have previously been linked to pediatric solid tumors, including OSA[65]. The 6p21 region has previously been identified to be amplified in OSAs. Furthermore, the 6p22 region contains a cluster of genes encoding histone proteins[66], five of which are included in the RFS signature, interesting because of histones’ role in epigenomic regulation. Very few pathways were associated with the CR signature, suggesting that broad methylation patterns reflect chemotherapy response in a pathway agnostic manner. Genes from the “14q32.31” locus were enriched in the CR signature. This is notable as we and others have previously reported miRNA expression from this imprinted region is associated with OSA biology and outcome[24, 25, 27]. Pathways enriched in the MetDx signature included “homophilic cell adhesion via plasma membrane adhesion molecules”, “cell fate commitment”, and multiple pathways relating to organ development. The MetDx signature was enriched for genes located on the “2q36.1” and “5q31” cytogenetic bands. We also analyzed the probes most significantly differentially methylated (p < 0.001, n = 7075 CpG sites, 1938 genes) between the global cluster groups (Fig. 1a) to understand the strongest differences in the broad methylation patterns observed. In this analysis, we again identified the “14q32.31” locus as the most enriched cytogenetic band, followed by the “1q22-23” region. The KEGG and BioCarta pathways most enriched in this subset of the global profile were “Cell adhesion molecules” and “Intrinsic Prothrombin Activation Pathway.” This is notable because of the interrelated roles these pathways are known to play in cancer invasion and migration[67, 68]. Specific to OSA, the coagulation factor F3, which enables the initiation of the blood coagulation cascade, has been shown to grant OSA metastatic competence and promotes disease progression[69-71]. Additionally, F3, four other coagulation factors, and two coagulation factor receptors were identified in the 95% most variantly methylated loci in the TARGET dataset.

We also identified genes previously reported to be involved in OSA biology in our outcome signatures. Methylation of ANGPT1, a gene important for angiogenesis and associated with a metastatic variant enhancer locus shown to promote OSA metastasis, was one of the methylation sites most strongly associated with RFS[69]. We also found TASOR2 and WWOX, both genes with SNPs previously identified as potential risk loci for OSA, in the RFS signature[72]. The RFS methylation signature also included loci encoding the miRNAs MIR211, which has been previously implicated in OSA prognosis[73], and MIR1247. The gene encoding telomerase (TERT) was identified in the CR signature. While about 60% of OSAs use the alternative lengthening of telomeres pathway instead of telomerase expression for telomere maintenance[74], a subset of tumors with increased TERT expression are known to be more aggressive[75].

### The RFS methylation markers may regulate a transcriptional program also prognostic of survival

After confirming the reproducibility of the outcome signature methylation patterns, we sought to study the outcome markers within their regulatory network. We used comprehensive multi-omic profiling data available in the TARGET dataset to assess the downstream consequences of methylation on gene transcription, assessed by RNA sequencing data available for the same samples. We studied the correlation between methylation levels of the outcome markers and transcription of the likely regulated genes based on genomic location, (details in Methods). We identified genes potentially associated with 71.7, 54.0, and 70.6% of methylation markers in the RFS, CR, and MetDx signatures, respectively (Table S1), and 51.8% of these expected correlations were significant (p <0.05). Additionally, these correlations were, on average, stronger than the outcome marker-transcription correlation where an interaction was not expected (average |r_expected_| = 0.412, average |r_other_| = 0.315). For the expected correlations, we found 30.5% were weak (|r| < 0.3), 56.7% were moderate (0.3 ≤ |r| ≤ 0.6), and 12.8% were strong (|r| > 0.6). When we considered the sign of the correlation coefficient and the signatures individually, we found transcriptional correlations with the RFS, CR, and MetDx markers were 54.1, 9.7, and 30.9% negative, respectively (Table S5). The prevalence of positive correlations could be partly explained by the large number of gene-body methylation sites in our signatures, which contained 75.2, 82.2, and 72.0% gene body (instead of promoter) sites in the RFS, CR, and MetDx signature, respectively.

While promoter associated methylation is known to inhibit transcription, gene-body methylation can be a mark of active transcription, albeit with a weaker connection[76]. Additionally, the small fraction of 3’ UTR site in the signature were all positively correlated with transcription, an observation recently reported in multiple adult cancer types[77]. These results suggest that many of the RFS methylation markers likely regulate transcription, but a smaller subset of the CR and MetDx are directly connected to downstream gene expression.

We also examined whether transcription at the methylation signature loci was associated with clinical outcomes. We compiled sets of genes (genesets) corresponding to each methylation signature based on the gene annotations of the methylation signature CpG sites. Test of association with the same clinical outcome predicted by the methylation signatures found showed the genesets to be significant using geneset enrichment analysis (GSA)[53] (Table 2). Of note, a large fraction of the CR methylation markers could not be annotated to a known gene because they were located in so called “gene deserts” which are enriched in the Open Sea genomic region[78], and the CR transcriptional geneset was much less strongly associated with outcome of the three genesets. As above, this suggests that a broad methylation cell state may be influencing response to chemotherapy, not necessarily mediated through direct transcriptional regulation of proximal genes by methylation. We also asked if the downstream transcriptional patterns classify samples similarly to the methylation profiles. We found this to be the case for the RFS signature with a moderately high degree of concordance (Cramer’s V = 0.443, p = 1×10^−4^). Furthermore, a survival difference was observed between two transcriptionally defined hierarchical clustering subgroups (Fig. 6), strengthening the connection between the RFS methylation markers and downstream transcriptional regulation. We did not find this concordance for the CR and MetDx signature (Fisher’s exact p = 0.315, and 0.402) further underscoring the concept that chemotherapy response is correlated with very broad methylation patterns rather than regulation of a few specific pathways via methylation.

**Table 2.** Gene set expression analysis (GSA) of genes regulated by the outcome methylation signatures. GSA results (LS / KS enrichment) analyze associations between collective transcription of the regulated gene sets and their respective clinical outcomes.

| **Transcription**<br>**signature** | **Number**<br>**of genes** | **GSA LS / KS**<br>**permutation p – value** |
| --- | --- | --- |
| RFS | 219 | $< 1 \times 10^{-5}$ |
| CR | 142 | 0.006 |
| MetDx | 220 | $< 1 \times 10^{-5}$ |

**Figure 6.**
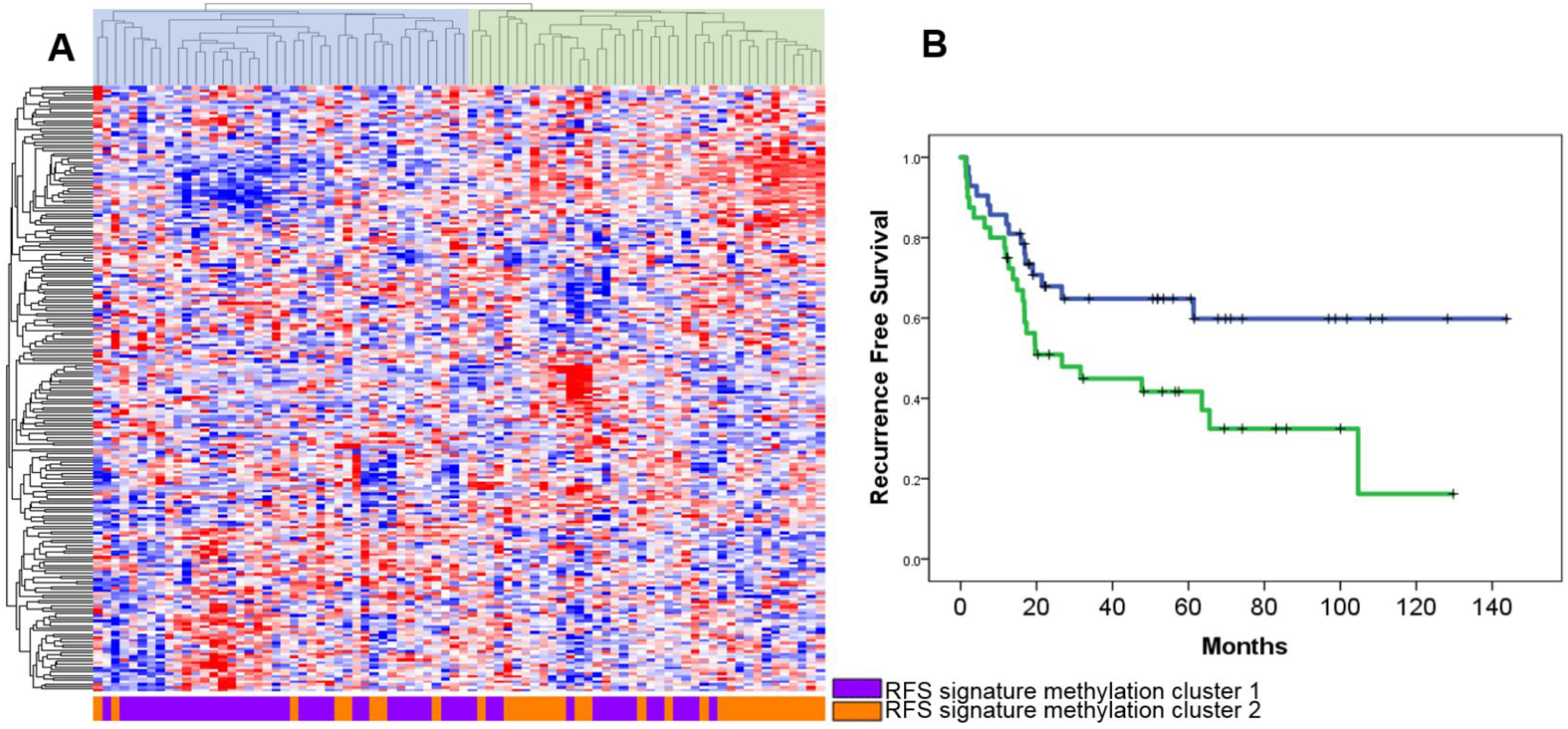
Genes regulated by the RFS methylation signature. **A)** Hierarchical clustering analysis using transcriptional patterns of the genes regulated by RFS methylation signature (2-group R-index = 0.78). Classification is similar to the respective RFS methylation cluster groups **(Fig. 3A)** (Cramer’s V = 0.443, p = 1 x 10^−4^). **B)** RFS analysis using the two main transcription-based cluster groups (Log Rank p value = 0.024, median RFS: NYR (Group 1, left) vs. 26.7 mo. (Group 2, right)).

To obtain insights into possible relevant pathways, we analyzed the transcriptional association of regulated downstream genes with outcome in the context of biologically informed predefined genesets also using GSA analysis and found transcription of many pathways associated with the RFS and MetDx outcomes (Table S6). “Response to Organic Cyclic Compounds”, “Cartilage Development”, and many pathways relating to neuronal activity and development were transcriptionally associated with RFS in the GSA analysis. Also, transcription of the “1p36” region, reported to be amplified in some OSAs, was associated with RFS[79]. Importantly, similar pathways were found enriched in the parent methylation RFS signature when we independently explored the methylation loci by functional annotation (Table S4), as was the case with the MetDx signature, which showed enrichment for “positive regulation of cell adhesion” and Gene Ontology (GO) terms “blood coagulation” and “hemostasis”, all pathways thought to play a known role in tumor metastasis[71, 80]. We found no transcriptional pathway associations with CR in the GSA analysis, further evidence that methylation may be predicting chemotherapy response independent of known regulatory pathways.

### *In vitro* correlates of the clinical methylation profiles

We used a public *in vitro* multi-omic and pharmacologic dataset (Genomics of Drug Sensitivity in Cancer (GDSC))[37-39] including 11 OSA cell lines (all from cases < 20 year old) to assess the *in vitro* correlates of the clinically derived methylation patterns as a prelude for future experimentation and therapeutic discovery. We compared the average β values of the global methylation and outcome signatures of the GDSC and TARGET datasets (Fig. 7A) and found that the methylation levels of the cell lines were similar to those of the clinical samples, albeit with a trend for CGI hypermethylation in the cell lines, as observed in other cancer types[81, 82]. We also assessed the similarity of *in vitro* methylation patterns using semi-supervised hierarchical clustering (Fig. 7B-D) and compared differential methylation between the two primary cluster groups using the same methodology used to compare the clinical datasets. Both global and outcome signature methylation patterns were similar between the *in vitro* and clinical datasets (Fig. 7 and Table S7). The RFS and MetDx signatures displayed a balance of hyper and hypo methylation, and the CR signature was almost entirely hypermethylated, similar to what was seen in the clinical datasets (Fig. 3 and 4),

**Figure 7.**
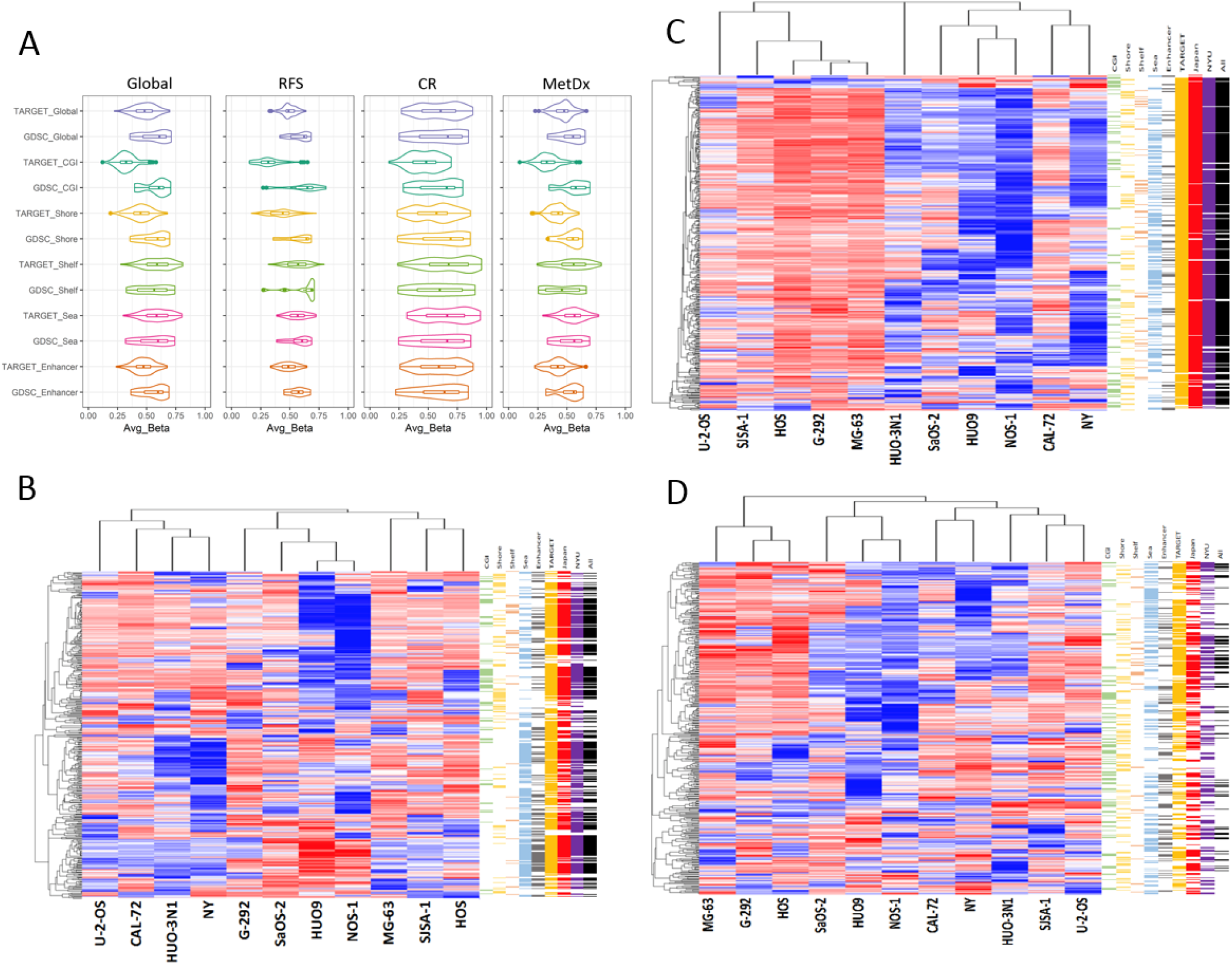
*In vitro* correlates of the methylation profiles. **A)** Plots displaying the average β value distributions of global methylation, outcome signatures, and genomic region subsets for the TARGET and GDSC datasets. **B, C**, and **D)** Semi-supervised hierarchical clustering of the 11 OSA cell lines in the GDSC dataset using the RFS **(B)**, CR **(C)**, and MetDx **(D)** outcome signatures. Genomic regions (first five color bars: CGI-green, Shore-yellow, Shelf-orange, Open Sea-blue, and Enhancer-grey) are shown to the right of each heatmap. CpG sites displaying concordant hypo / hyper methylation patterns in the in-vitro dataset relative to each of the clinical datasets are also annotated (last four color bars: TARGET-orange, JNCCRI-red, NY-purple, and all three clinical datasets-black) are annotated to the right of each heatmap. Detailed concordance results are presented in **Table S6**.

Cell line aggressiveness metrics, including proliferation, invasion, migration, colony forming ability, and tumorigenicity, were previously reported for 22 OSA cell lines[83], 7 of which were also profiled in the GDSC dataset methylation dataset. We tested the TARGET defined methylation patterns in relation to these cell line metrics. While we found that a relatively small fraction of individual CpG sites in our RFS and MetDx signatures correlated with *in vitro* aggressiveness metrics, permutation testing indicated the RFS signature is enriched for CpG sites associated with invasion, migration, and proliferation, and the MetDx signature is enriched for sites associated with invasion and migration (Table S8).

The same GDSC datasets also provided *in vitro* drug response testing for standard chemotherapeutic agents used in OSA (MAP: methotrexate, Adriamycin (doxorubicin), and cisplatin). We found a small number of individual CpG sites associated with response to standard chemotherapeutic agents *in vitro* (Table S9). However, poor response to doxorubicin, potentially the single most active drug used to treat OSA, was correlated with hypermethylation of the vast majority of associated CpG sites, similar to the large hypermethylation - poor chemotherapy response association observed in the TARGET dataset. The three standard drugs (MAP) were not tested in combination *in vitro*, (as they are always used clinically). Thus, we created a surrogate “MAP response” variable by scaling each cell line’s response to each drug to a value between 0 and 1, where 0 and 1 were assigned to the cell lines least and most resistant to the drug, respectively, and then averaged the resulting scores. We observed hypermethylation was positively correlated with this variable, again indicating the chemoresponse patterns associated with methylation *in vivo* are also observed *in vitro*.

### Large-scale integrative pharmacogenomic analysis identifies drugs with potential activity in OSA

We sought to test if the RFS signature can be a helpful tool to screen for novel drugs and drug repurposing. At this point, there is no analytical pipeline optimized for use of methylomic patterns for pharmacogenomic analysis. Therefore, we used the transcription patterns as a surrogate. We did not use the CR signature as we did not find a clear transcriptional correlate for the extremely broad profiles of chemoresponse. We leveraged our comprehensive pharmacogenomic analytical platform PharmacoGx through the PharmacoDB interface[55, 56], which we have previously used for drug discovery[27, 84], in a “pan cancer” discovery process increasing sensitivity for markers of drug response.

Among the genes annotated to the RFS methylation signature, we sub-selected 47 with at least a moderately strong correlation with the RFS methylation markers (|r| > 0.3, p < 0.001) to increase confidence that these genes are regulated by methylation. We tested these 47 genes for drug response association in a pan-cancer analysis using the 1,691 cancer cell lines with the 759 compounds included in the database. Initial drug selection based on a regression analysis identified 61 drugs (further details in Methods). Then we focused our analysis on the subset of 11 OSA cell lines to increase specificity and eliminated 1/3 of drugs with the least variant IC50 in the OSA cell lines. We selected drugs for which the direction (+ or – regression coefficient) of the gene-drug association observed in the pan-cancer analysis was the same as in the OSA specific analysis for at least two thirds of the significant genes, and to increase clinical impact, we considered drugs more potent than cisplatin in at least 2 of the 11 OSA cell lines. The resulting list of 18 drugs is presented in Table 3. We then used the *in vitro* methylomic dataset described above (GDSC) to search for more specific evidence of relationship between drug response and methylation, and we found that for at least 5 of the drugs in Table 3, the RFS signature showed a stronger association with response than 80% of random CpG site sets of equivalent size (details in Methods).

**Table 3.**
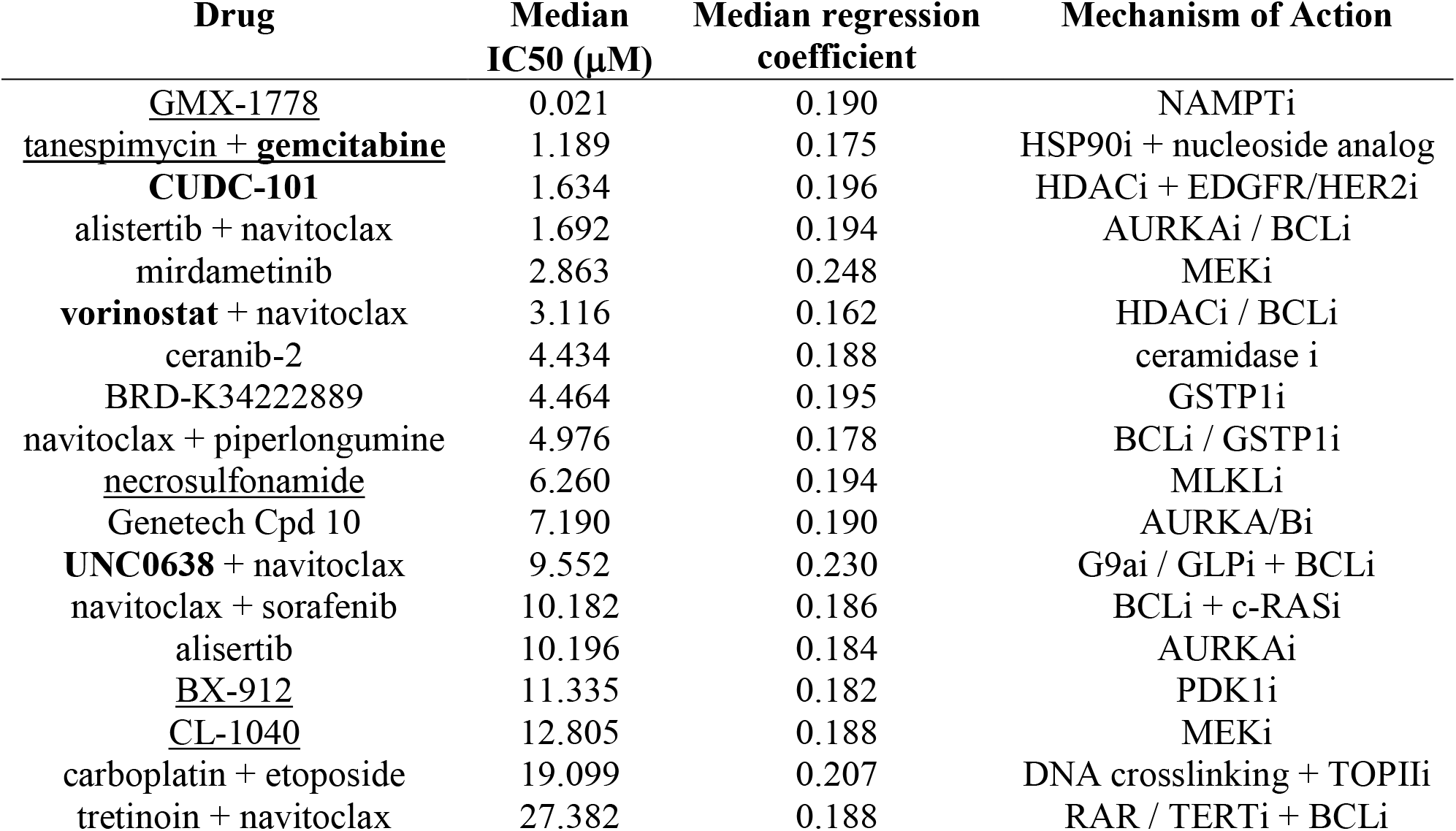
Pharmaceuticals for which PharmacoDB analysis reveals a predictive drug response association with the transcription of genes regulated by the RFS methylation signature. Median IC50 values are obtained across all OSA cell lines in the GDSC dataset. The median regression coefficient is calculated from the significant gene – drug interactions used to identify the drugs. Bolded drugs act through epigenetic mechanisms. Underlined drugs have more strong response correlations (|r| > 0.6) with the RFS methylation signature than at least 80% of the randomly generated CpG site sets in OS cell lines.

Our pipeline uncovered both novel and clinically developed drugs. Four identified drugs act through epigenetic mechanisms. CUDC-101 and vorinostat inhibit histone deacetylases. UNC0638 inhibits histone methylation through G9a and GLP. G9a has previously been linked to response to cisplatin[85]. Furthermore, UNC0638 acts at several miRNA encoding loci across the genome[86]. It has been reported that DNA and histone methyltransferase inhibitors preferentially affect expression of genes silenced in cancer, whereas HDAC inhibitors alter transcription of about 1/3 of the genome[34]. We also identified tretinoin, which is known to inhibit telomerase[87]. As previously mentioned, while many OSAs are thought to preserve telomere length through the alternative lengthening of telomeres pathway, an aggressive subset of tumors is known to aberrantly express telomerase[75]. Furthermore, methylation of the telomerase (TERT) genomic locus was found to be associated with response to chemotherapy (Table S1). The nucleoside analog gemcitabine, which inhibits DNA methyltransferases (DNMTs) in a similar manner to demethylating agents azacitidine and decitabine was also in thelist[88]. In a prespecified hypothesis-based analysis, we tested if azacytidine was associated with the RFS signature (p<0.001) using somewhat relaxed regression coefficient criterion (coefficient >0.1) and found four associations between decitabine response and the RFS signature in the pan cancer analysis. Only one OSA cell line had available decitabine IC50 data precluding additional *in vitro* analysis. While more drugs targeting DNA methylation via DNMT inhibition and other mechanisms such as IDH1/2 inhibition have been developed, these drugs were not included in the PharmacoDB dataset.

Given the recent interest in cabozantinib and pazopanib in OSA, we also performed a targeted test for association between the RFS signature and response to these drugs[89-94]. In a prespecified hypothesis-based analysis (p<0.001, coefficient >0.1), we found that one and two of the RFS signature genes were associated with cabozantinib and pazopanib response, respectfully. When we evaluated the response associations specifically in the OSA cell lines, we found that the direction of the gene-response association for cabozantinib, but not those for pazopanib, were concordant with the pan-cancer analysis. Finally, in the GDSC methylomic data, we found more methylation sites strongly correlating (|r| > 0.6) with response than all 10 randomly generated CpG sets.

We compared the list of drugs in Table 3 to those we had previously discovered using a very similar miRNA gene target-based pharmacogenomics analysis using the same bioinformatic platform[27]. Only one drug (mirdametinib (PD-0325901)) was identified in both analyses suggesting methylation and microRNA molecular patterns offer non-overlapping insight into the disease and drug discovery.

## DISCUSSION

Despite ongoing efforts, there has been no meaningful advancement in OSA therapies or biomarkers for the last three decades. Conventional genetic studies have not uncovered actionable drug targets, and the only accepted prognostic factor is pathologically assessed tumor necrosis following standard neoadjuvant therapy. This did not lead to improvement in survival when used to stratify patients in a recent large randomized international trial (EURAMOS)[6, 22, 23] and recent consensus has emerged that there is pressing need for new robust biomarkers that better align with biologic and clinical heterogeneity in this tumor[95, 96]. The state of DNA methylation is often disrupted in cancer in many major ways[97]. Furthermore, DNA methylation has been shown to carry prognostic information and define disease subtypes with different treatment response in other cancers[98-100]. In OSA, prior reports have described single gene or locus methylation affecting progression or treatment response[30, 31]. A pilot clinical study of 15 patients suggested that the genome-wide DNA methylation state of pre-treatment OSA tumors is prognostic of outcome[36].

We analyzed the recent multi-omic NCI TARGET OSA dataset, representing the largest (N=83) methylation profiling study conducted thus far in this relatively rare, largely pediatric disease. Genome-wide analysis revealed that primary tumor methylation patterns were strongly associated with patient outcomes. Most striking was the large genome-wide difference in methylation state between tumors which did and did not respond to standard therapy, with the relatively hypomethylated tumors responding better to chemotherapy than the hypermethylated tumors. This was found by both unsupervised clustering, and a supervised approach whereby the methylation level of genome-wide CpG sites was simply averaged for each sample, suggesting that the clinically relevant DNA methylation patterns are present across a large fraction of the genome, and not limited to a few genomic loci. Notably, broad genomic hypomethylation is known to cause genomic instability and increase tumor immunogenicity. This is especially interesting in light of recent immunogenomic findings suggesting that OSA tumors with optimal chemoresponse have less stable genomes and higher activation of immune response than those with suboptimal response[101].

Methylation patterns were also found to predict long-term survival. Unsupervised clustering of sample groups revealed that the same genome-wide methylation patterns were also prognostic of RFS, when stratified for the presence of metastasis at the time of diagnosis. When we focused on specific regions of the genome based on CpG density, we found that methylation of CGIs was most strongly prognostic of long-term outcome, as has been described in other cancer types[98]. Intriguingly, we found that CGI methylation patterns within a given risk group were heterogenous, unlike the CpG sparse genomic regions (Open Sea) which were more uniformly hyper or hypomethylated in a given methylation subgroup. It is, thus, conceivable that CGI methylation contributes to locus specific regulation mediating gene transcriptional programs, whereas Open Sea regions represent a biologically meaningful broader genomic state.

In addition, a supervised RFS prognostic signature was discovered with strong FDR corrected statistical significance, although a model based on this signature could not be independently validated given the current lack of other available large clinical methylomic datasets. Analysis of this RFS signature revealed a subpopulation of CpG sites where the CGIs were primarily hypomethylated, and Open Sea sites were primarily hypermethylated, in a poor prognosis subgroup. Further support for the notion that different regions offer non-overlapping prognostic information was obtained through multivariate analysis of the RFS signature while controlling for response to chemotherapy. CpG sites in CGIs were depleted in the RFS signature but enriched when only considering the subset of sites predictive of outcome independent of chemoresponse. and even suggested an inverse methylation-outcome relationship (with hypermethylation associating with better outcome) compared to the genome wide pattern. This contrasted with the chemoresponse (CR) signature, which was nearly ubiquitously hypermethylated in the poor response group, except for a very small subset of CGI and Shore sites. The cancer genome is generally known to gain methylation at CGIs and lose methylation in Open Sea regions[61], thus it is interesting to observe that our signatures predict that tumors with methylation patterns more reminiscent of those of normal tissue may be more aggressive or less responsive to standard chemotherapy.

While have previously reported that miRNA patterns are prognostic of survival we have found limited evidence to date for a robust and reproducible miRNA signature associated with chemotherapy response or metastatic disease at the time of diagnoses. Our results suggest that the methylation signatures are offering independent, and more powerful chemoresponse prediction than miRNA transcription. In this respect, multivariate analysis also demonstrated that the association of the RFS methylation signature was independent of that of the previously published miRNA survival signatures. Thus, these different omics patterns can be potentially used in a complementary manner to identify clinically relevant tumor subtypes in OSA.

Analysis of two independent publicly available but relatively small clinical datasets allowed us to demonstrate the reproducibility of methylation molecular profile. While validation of the outcome signatures could not be performed because the public datasets lacked survival or chemoresponse information, we report that the molecular information carried in each dataset could be used to group samples with similar methylation patterns as observed in the large TARGET cohort, suggesting the methylation profiles may indeed define reproducible molecular subtypes. Furthermore, our results extend upon those from the previous study of the prognostic relevance of DNA methylation, which profiled 15 samples using a HELP-tagging assay, which found increased genome-wide DNA methylation in primary tumors of patients that eventually relapsed compared to those that remained in remission[36]. This parallels our observation from genome-wide clustering analysis where the hypermethylated subgroup displayed worse RFS. Likewise, both studies identify a preponderance of either hyper or hypomethylation in Open Sea regions, a more even ratio of hyper to hypomethylation at CGIs in individual tumors, and relative hypermethylation of enhancer regions associated with high risk for recurrence. Beyond these findings, our study is the first to show a striking powerful prediction of chemoresponse by broad methylation profiles and signatures.

When analyzing the relationship between all outcome signatures CpG sites and known genes, we observed that many sites were not annotated to a known gene (intergenic), and those that were intragenic often fell within gene bodies rather than promoter regions. Similarly, the previous pilot study observed an overrepresentation of CpG sites related to chemoresponse in intergenic regions, and an underrepresentation of these sites in promoter loci.

Gene expression analysis of the same samples profiled in the TARGET methylation cohort revealed a possible connection between the methylation profiles and transcription from these genomic loci. This connection was quite robust for the RFS signature, where methylation and downstream transcription profiles were found to carry similar prognostic information, suggesting methylation may be activating or repressing key genes. Connection between the CR signature and transcription was weak, suggesting that the larger epigenomic cellular state may be a better marker for response to standard therapy than a methylation effect on a few specific critical signaling pathways.

We used the down transcriptional surrogate of the RFS methylation signature to perform pharmacogenomic drug discovery analysis using a novel bioinformatic pipeline[55, 56] and identified a set of drugs for which methylation patterns might serve as response biomarkers. These drugs were almost entirely non-overlapping with a set of candidate drugs from a similar analysis we previously reported which used the gene targets of prognostic miRNA profiles, suggesting again that methylation and miRNA patterns offer non-redundant clinically applicability[27]. Several of these drugs act through epigenomic mechanisms, especially modulation of post-translational histone modifications. Histone modifications influence and can be influenced by the DNA methylation state of the underlying locus and provide modifiable regulation of gene expression, an active area of research in other solid and hematological malignancies[102]. Notably, the HDAC4 gene (the protein product of which deacetylates core histones) and the gene of its binding partner MEF2C[103], were found in the RFS signature, which also included five replication dependent core (H2A, H2B, H3, and H4) histones from the H1 histone cluster on chromosome 6[104]. Three of the histone genes identified in the signature (HIST1H2BK, HIST1H3J, and HIST1H4I) have been shown to have dysregulated transcription in other tumors[105]. The HIST1H4I gene has been shown to be differentially methylated in parathyroid tumors compared to normal tissue[106]. Mutations of H3 histones are highly prevalent in other bone tumors such as giant cell tumors of bone, chondroblastomas, and chondrosarcomas and are also found in pediatric brain tumors and were recently also identified in a small subset of OSAs, present exclusively in adult patients[107-112]. These H3 mutant tumors displayed differential methylation patterns compared to H3 wildtype OSAs, and one of the two most differentially methylated sites was in the histone micro-cluster included in the RFS signature[109].

Demethylating drugs have shown pre-clinical efficacy in OSA[30, 31]. Furthermore, standard chemotherapy includes cisplatin and doxorubicin, both of which induce direct DNA damage. Hypermethylation may protect against these insults or even physically disrupt their mechanisms by limiting chromatin accessibility or stabilizing the genome. Moreover, epigenetic targeting has been shown to potentiate chemotherapy effect, and a recent study reported that a DNA methyltransferase inhibitor synergized with both doxorubicin and cisplatin in OSA cell lines[113, 114]. Furthermore, HDAC inhibitors synergize with doxorubicin in short term cultures derived from orthotopic patient derived xenograft models[115]. Finally, our finding that the EPIMMUNE signature, recently reported to predict immunotherapy response in lung cancer, is detectable in a subset of osteosarcoma samples merits further dedicated investigation. Thus, our study implies that epigenetic therapies hold promise for synergistic combination with standard chemotherapy and be especially useful in the subset of hypermethylated tumors expected to respond poorly to chemotherapy

Limitations of our study include the lack of available survival data in the independent public datasets, which allowed us to reproduce the molecular profiles, but not to fully validate the prognostic associations, though the statistical signal in the large TARGET dataset is extremely strong and appropriately controlled for high dimensional analysis. Additionally, we could not evaluate a possible association with histologic subtypes given the lack of such information in any of the public datasets that were available. Additional validation, optimization, and translation to a more targeted clinically applicable assay will be needed prior to the clinical application of the methylation signatures described in this report. Standard clinicopathologic variables alone have thus far not been adequate as markers for improved or properly tailored therapies for OSA, highlighted by the failure of the recent EURAMOS trial[6, 23], which used pathologically assessed tumor necrosis to stratify patients for alternate or intensified therapy. DNA methylation signatures herein, may ultimately offer a powerful and biologically informed method to complement clinical prognostic factors for therapeutic stratification.

Additionally, recent work has shown that tumor DNA methylation patterns are preserved in formalin fixed paraffin embedded (FFPE) tissue facilitating the study and application of methylation markers in rare tumors like OSA[116, 117], as well as in the blood stream[118, 119], such that minimally invasive methylation assays assisting clinical management of OSA may ultimately become feasible.

## CONCLUSIONS

We present the largest epigenomic study of osteosarcoma to date. We discovered that genome-wide methylation patterns define OSA patient risk subgroups with very different survival and response to standard therapy. We also defined CpG site specific methylation signatures predictive of clinical outcomes. The discovered methylation patterns are reproducible in independent clinical and in vitro datasets. Furthermore, we found that the methylation patterns are associated with transcriptional networks, suggesting they may characterize molecular disease subtypes. These findings open the possibility for stratifying patients for alternate therapies based on their methylation profile, and potentially suggest targeting epigenetic aberrations may be a useful treatment strategy in this disease.

## Data Availability

The datasets analyzed in this study are available at the TARGET data matrix (https://ocg.cancer.gov/programs/target/data-matrix), the Gene Expression Omnibus repository (GSE125645, GSE97529, and GSE68379), ArrayExpress (E-MTAB-3610), and the PharmacoDB database (https://pharmacodb.pmgenomics.ca/).

## ABBREVIATIONS

OSA: osteosarcoma
TARGET: Therapeutically Applicable Research to Generate Effective Treatments
RFS: Recurrence Free Survival
CR: Chemotherapy Response
MetDx: Metastatic disease at the time of Diagnosis
miRNA: microRNA
JNCCRI: Japanese National Cancer Center Research Institute
NY: New York
GDSC: Genomics of Drug Sensitivity in Cancer
KM: Kaplan-Meier
FDR: False Discovery Rate
OR: Odds Ratio
GO: Gene Ontology
KEGG: Kyoto Encyclopedia of Genes and Genomes
CGI: CpG Island
OS: Overall Survival
MAP: Methotrexate, Adriamycin, and Cisplatin

## DECLARATIONS

### Ethics approval and consent to participate

All data analyzed was publicly available and experiments were approved by the presiding Institutional Review Board where each dataset was generated. All data analyzed was generated by the NCI TARGET initiative (https://ocg.cancer.gov/programs/target) or has been previously published [30, 35].

### Consent for publication

Not applicable

## Authors’ contributions

DS, MA, and CL conceived the study, CL performed experiments and analysis, CL, EN, AK, SLC, GC, BHK, and DS wrote the manuscript. All authors contributed to the interpretation of the results and provided critical feedback on the manuscript. All authors read and approved the final manuscript.

## Acknowledgements

The results published here are in part based upon data generated by the Therapeutically Applicable Research to Generate Effective Treatments (https://ocg.cancer.gov/programs/target) initiative, phs000218. The data used for this analysis are available at https://portal.gdc.cancer.gov/projects.

## Supplementary table and figure legends

**Table S1**. Methylation of CpG sites statistically associated with specific outcomes. Associations with RFS were assessed using Cox Regression analysis and associations with CR and MetDx were assessed using a two-sample t-test. Both a nominal p < 0.05 and a Benjamini-Hochberg False Discovery Rate (FDR) corrected p < 0.1 were used to define significance for RFS and CR associations. An uncorrected cutoff of p < 0.05 was used to define significance for MetDx associations. To define signatures specifically associated with each outcome CpG sites associated with multiple outcomes were excluded from the signatures. Given the fact that the lists were very unequal in size we arbitrarily defined the signatures as the list of top markers equal in size to the smallest list of outcomes associated methylation sites (374 sites). **See separate txt file**.

**Table S2**. Genomic region enrichment in the methylation profiles. Enrichment (green) or depletion (red) significance as determined by the hypergeometric test.

**Table S3**. Differential methylation analysis of the global methylation patterns and three outcome signatures in three independent clinical datasets. Fraction of differentially methylated (p < 0.05) CpG sites between the two primary cluster groups that were concordantly hypo / hyper methylated in the independent datasets relative to the TARGET dataset is shown **(Fig. 4)**.

**Table S4**. Functional annotations using the genes regulated by the outcome methylation profiles. Pathways and terms with an FDR corrected p value < 0.1 were considered significant. **See separate txt file**.

**Table S5**. Correlation between the methylation outcome signatures and gene transcription. CpG methylation (M values) and gene transcription were analyzed by Pearson’s correlation for each gene proximal to the CpG location based on array annotations. Significant correlations (p < 0.05) with |r| < 0.3 were considered weak, with 0.3 ≤ |r| ≤ 0.6 were considered intermediate, and with |r| > 0.6 were considered strong, suggesting higher likelihood for mechanistic specificity.

**Table S6**. Geneset enrichment (GSA analysis) using the genes regulated by the outcome methylation profiles. Pathways and terms with a LS or KS permutation p value < 0.05 and a LS and KS permutation p value < 0.1 were considered significant.

**Table S7**. Differential analysis of the global methylation patterns and three outcome signatures in the clinical and *in vitro* datasets. The fraction of differentially methylated (p < 0.05) CpG sites between the two primary cluster groups **(Fig. 7)** that were concordantly hypo / hyper methylated between the *in vitro* and each clinical dataset are shown.

**Table S8**. Correlations between methylation of the outcome signature CpG sites and *in vitro* aggressiveness metrics. The fraction of probes strongly (|r| < 0.6) correlated with the aggressiveness metrics is presented.

**Table S9**. Correlations between methylation of the outcome signature CpG sites and *in vitro* response to standard chemotherapy. The fraction of probes strongly (|r| < 0.6) correlated with standard chemotherapeutics is presented. MAP was not tested as a combination treatment, so combined MAP response was assessed by combining methotrexate, doxorubicin, and cisplatin response metrics.

**Figure S1**. Survival analysis of the two main hierarchical clustering groups generated using each of the genomic regions when stratified for metastasis at the time of diagnosis. CGI region: RFS (**A**, pooled p = 0.002) OS (**B**, pooled p = 0.001). Shore region: RFS (**C**, pooled p = 0.036), OS (**D**, pooled p = 0.005). Shelf region: RFS (**E**, pooled p = 0.006), OS (**F**, pooled p = 0.044). Open Sea region: RFS (**G**, pooled p = 0.011), OS (**H**, pooled p = 0.002). Enhancer region: RFS (**I**, pooled p = 0.003), OS (**J**, pooled p = 0.008).

**Figure S2**. RFS analysis of patient risk groups generated with methylation of single CpG site from each analyzed genomic region. Patient risk groups were generated by methylation level (median split and terciles). **A)** CGI region CpG site cg19848683 (MECOM), 2-group log-rank p value = 0.005, 3-group log-rank p value = 0.003. **B)** Shore region, CpG site cg04461028 (HDAC4), 2-group log-rank p value < 0.001, 3-group log-rank p value = 0.004. **C)** Shelf region, CpG site cg06835212 (MEF2C) 2-group log-rank p value = 0.002, 3-group log-rank p value < 0.001. **D)** Open Sea region, CpG site cg15595627 (ANGPT1), 2-group log-rank p value < 0.001, 3-group log-rank p value < 0.001. **E)** Enhancer region, CpG site cg12506775 (TASOR2), 2-group log-rank p value < 0.001, 3-group log-rank p value < 0.001.

**Figure S3**. Analysis of the EPIMMUNE lung cancer methylation signature. Hierarchical clustering and the heatmap were generated using M values of the 128 EPIMMUNE CpG sites mapped to the TARGET osteosarcoma dataset (two group cluster stability R index = 0.745). Green highlighted samples have a methylation profile predictive of optimal immune checkpoint inhibitor response in the EPIMMUNE discovery cohort. Sites hyper- (red) or hypo- (blue) methylated in immune checkpoint inhibitor responders in the EPIMMUNE study discovery cohort are displayed to the right of the heatmap (leftmost color bar). Genomic regions (middle color bars) and sites passing the 95% variance filtering criteria used for discovery analysis in the TARGET dataset (rightmost color bar) are also displayed. Color bars below the heatmap annotate TARGET osteosarcoma sample response to chemotherapy, and cluster group membership from the genome-wide clustering analysis **(Fig. 1A)**.

