## Supplementary Materials for "Genome-wide DNA methylation patterns reveal clinically relevant predictive and prognostic subtypes in osteosarcoma"

**Description of Supplementary Files**

**Table S1.** Methylation of CpG sites statistically associated with specific outcomes. Included as a separate file.

**Table S2.** Genomic region enrichment in the methylation profiles.

**Table S3.** Differential methylation analysis of the global methylation patterns and three outcome signatures in three independent clinical datasets.

**Table S4.** Functional annotations using the genes regulated by the outcome methylation profiles. Included as a separate file.

**Table S5.** Correlation between the methylation outcome signatures and gene transcription.

**Table S6.** Geneset enrichment (GSA analysis) using the genes regulated by the outcome methylation profiles.

**Table S7.** Differential analysis of the global methylation patterns and three outcome signatures in the clinical and *in vitro* datasets.

**Table S8.** Correlations between methylation of the outcome signature CpG sites and *in vitro* aggressiveness metrics.

**Table S9.** Correlations between methylation of the outcome signature CpG sites and *in vitro* response to standard chemotherapy.

**Figure S1.** Survival analysis of the two main hierarchical clustering groups generated using each of the genomic regions when stratified for metastasis at the time of diagnosis.

**Figure S2.** RFS analysis of patient risk groups generated with methylation of single CpG site from each analyzed genomic region.

**Figure S3.** Analysis of the EPIMMUNE lung cancer methylation signature.

**Table S1.** Methylation of CpG sites statistically associated with specific outcomes. Associations with RFS were assessed using Cox Regression analysis and associations with CR and MetDx were assessed using a two-sample t-test. Both a nominal $p < 0.05$ and a Benjamini-Hochberg False Discovery Rate (FDR) corrected $p < 0.1$ were used to define significance for RFS and CR associations. An uncorrected cutoff of $p < 0.05$ was used to define significance for MetDx associations. To define signatures specifically associated with each outcome CpG sites associated with multiple outcomes were excluded from the signatures. Given the fact that the lists were very unequal in size we arbitrarily defined the signatures as the list of top markers equal in size to the smallest list of outcomes associated methylation sites (374 sites). **See separate txt file.**

**Table S2.** Genomic region enrichment in the methylation profiles. Enrichment (green) or depletion (red) significance as determined by the hypergeometric test.

| Region | Global (95% most variantly methylated sites | RFS signature | CR signature | MetDx signature |
|---|---|---|---|---|
| **CGI** | $1.91x10^{-53}$ | $5.23x10^{-6}$ | $1.39x10^{-4}$ | 0.178 |
| **CGI $_{promoter\ associated}$** | $1.19x10^{-242}$ | $8.97x10^{-3}$ | $8.06x10^{-4}$ | $8.97x10^{-3}$ |
| **Shore** | $1.03x10^{-3}$ | 0.0179 | 0.166 | 0.0915 |
| **Shelf** | $1.38x10^{-11}$ | 0.522 | 0.020 | 0.371 |
| **Open Sea** | $7.31x10^{-56}$ | $4.48x10^{-9}$ | $1.24x10^{-3}$ | 0.471 |
| **Enhancer** | $1.33x10^{-51}$ | $1.63x10^{-6}$ | $5.83x10^{-3}$ | 0.319 |
| **Genic** | $1.03x10^{-281}$ | $2.59x10^{-3}$ | $4.94x10^{-6}$ | $9.62x10^{-3}$ |

**Table S3.** Differential methylation analysis of the global methylation patterns and three outcome signatures in three independent clinical datasets. Fraction of differentially methylated ($p < 0.05$) CpG sites between the two primary cluster groups that were concordantly hypo- / hyper- methylated in the independent datasets relative to the TARGET dataset is shown **(Fig. 4)**.

| Profile | TARGET v. JNCCRI (%) | TARGET v. NY (%) | JNCCRI v. NY (%) |
|---|---|---|---|
| Global | 71.41 | 55.38 | 74.81 |
| RFS | 99.00 | 98.78 | 100 |
| CR | 100 | 100 | 100 |
| MetDx | 82.46 | 72.22 | 92.31 |

**Table S4.** Functional annotations using the genes regulated by the outcome methylation profiles. Pathways and terms with an FDR corrected p value $< 0.1$ were considered significant. **See separate txt file.**

**Table S5.** Correlation between the methylation outcome signatures and gene transcription. CpG methylation (M values) and gene transcription were analyzed by Pearson's correlation for each gene proximal to the CpG location based on array annotations. Significant correlations ($p < 0.05$) with $|r| < 0.3$ were considered weak, with $0.3 \leq |r| \leq 0.6$ were considered intermediate, and with $|r| > 0.6$ were considered strong, suggesting higher likelihood for mechanistic specificity.

| Signature | Location | Probe – gene pairs | all significant | | weak | | intermediate | | strong | |
|---|---|---|---|---|---|---|---|---|---|---|
| | | | positive | negative | positive | negative | positive | negative | positive | negative |
| RFS | All | 282 | 61 | 72 | 27 | 25 | 26 | 41 | 8 | 6 |
| | TSS1500 | 41 | 5 | 11 | 3 | 4 | 2 | 5 | 0 | 2 |
| | TSS200 | 29 | 9 | 12 | 6 | 3 | 3 | 9 | 0 | 0 |
| | 5'UTR | 33 | 4 | 8 | 0 | 4 | 3 | 4 | 1 | 0 |
| | 1st Exon | 13 | 4 | 4 | 4 | 2 | 0 | 2 | 0 | 0 |
| | Body | 148 | 29 | 37 | 8 | 12 | 15 | 21 | 6 | 4 |
| | 3'UTR | 18 | 10 | 0 | 6 | 0 | 3 | 0 | 1 | 0 |
| CR | All | 208 | 84 | 9 | 20 | 8 | 57 | 0 | 7 | 1 |
| | TSS1500 | 28 | 9 | 3 | 2 | 3 | 6 | 0 | 1 | 0 |
| | TSS200 | 9 | 1 | 0 | 0 | 0 | 0 | 0 | 1 | 0 |
| | 5'UTR | 18 | 4 | 4 | 0 | 3 | 3 | 0 | 1 | 1 |
| | 1st Exon | 10 | 3 | 0 | 2 | 0 | 1 | 0 | 0 | 0 |
| | Body | 140 | 66 | 2 | 16 | 2 | 47 | 0 | 3 | 0 |
| | 3'UTR | 3 | 1 | 0 | 0 | 0 | 0 | 0 | 1 | 0 |
| MetDx | All | 296 | 125 | 56 | 24 | 20 | 74 | 33 | 27 | 3 |
| | TSS1500 | 65 | 14 | 14 | 4 | 7 | 10 | 6 | 0 | 1 |
| | TSS200 | 18 | 2 | 7 | 1 | 2 | 0 | 5 | 1 | 0 |
| | 5'UTR | 26 | 11 | 9 | 1 | 2 | 3 | 7 | 7 | 0 |
| | 1st Exon | 12 | 1 | 9 | 1 | 2 | 0 | 5 | 0 | 2 |
| | Body | 165 | 90 | 17 | 15 | 7 | 59 | 10 | 16 | 0 |
| | 3'UTR | 10 | 7 | 0 | 2 | 0 | 2 | 0 | 3 | 0 |

**Table S6.** Geneset enrichment (GSA analysis) using the genes regulated by the outcome methylation profiles. Pathways and terms with a LS or KS permutation p value < 0.05 and a LS and KS permutation p value < 0.1 were considered significant.

| Signature | Term source | Term | Genes | LS perm. p | KS perm. p |
|---|---|---|---|---|---|
| RFS | GO | regulation of synaptic plasticity | 6 | 0.001 | 0.004 |
| RFS | GO | learning or memory | 5 | 0.002 | 0.008 |
| RFS | GO | cognition | 5 | 0.002 | 0.008 |
| RFS | GO | modulation of chemical synaptic transmission | 7 | 0.002 | 0.017 |
| RFS | GO | regulation of trans-synaptic signaling | 7 | 0.002 | 0.017 |
| RFS | GO | regulation of signaling receptor activity | 8 | 0.003 | 0.056 |
| RFS | GO | cartilage development | 7 | 0.005 | 0.079 |
| RFS | GO | chemical synaptic transmission | 17 | 0.005 | 0.047 |
| RFS | GO | anterograde trans-synaptic signaling | 17 | 0.005 | 0.047 |
| RFS | GO | synaptic signaling | 17 | 0.005 | 0.047 |
| RFS | GO | trans-synaptic signaling | 17 | 0.005 | 0.047 |
| RFS | GO | cellular response to organic cyclic compound | 6 | 0.008 | 0.001 |
| RFS | GO | positive regulation of nervous system development | 11 | 0.009 | 0.002 |
| RFS | GO | positive regulation of neuron differentiation | 9 | 0.009 | 0.001 |
| RFS | GO | positive regulation of neurogenesis | 9 | 0.009 | 0.001 |
| RFS | GO | regulation of neuron differentiation | 14 | 0.012 | 0.014 |
| RFS | GO | receptor regulator activity | 6 | 0.015 | 0.077 |
| RFS | GO | receptor ligand activity | 6 | 0.015 | 0.077 |
| RFS | GO | cellular response to endogenous stimulus | 21 | 0.018 | 0.012 |
| RFS | GO | endoplasmic reticulum part | 15 | 0.019 | 0.089 |
| RFS | GO | dephosphorylation | 5 | 0.019 | 0.019 |
| RFS | GO | regulation of neuron projection development | 10 | 0.021 | 0.034 |
| RFS | GO | lipid biosynthetic process | 7 | 0.022 | 0.040 |
| RFS | GO | regulation of neurogenesis | 17 | 0.023 | 0.034 |
| RFS | GO | neuron projection development | 17 | 0.024 | 0.035 |
| RFS | GO | response to organic cyclic compound | 8 | 0.024 | 0.033 |
| RFS | GO | response to radiation | 8 | 0.028 | 0.057 |
| RFS | GO | neural retina development | 6 | 0.032 | 0.047 |
| RFS | GO | response to nitrogen compound | 12 | 0.040 | 0.092 |
| RFS | GO | cellular response to radiation | 5 | 0.040 | 0.095 |
| RFS | GO | regulation of nervous system development | 19 | 0.046 | 0.046 |
| RFS | GO | response to endogenous stimulus | 25 | 0.047 | 0.029 |
| RFS | GO | positive regulation of cell development | 13 | 0.049 | 0.027 |
| RFS | MSigDB_positional | chr1p36 | 5 | 0.028 | 0.016 |
| RFS | MSigDB_oncogenic | LEF1_UP.V1_DN | 5 | 0.014 | 0.090 |
| RFS | MSigDB_miRNA | CTTTGCA_MIR527 | 5 | 0.017 | 0.029 |
| RFS | MSigDB_miRNA | TAGCTTT_MIR9 | 5 | 0.035 | 0.061 |
| RFS | MSigDB_miRNA | CTTTGTA_MIR524 | 5 | 0.039 | 0.050 |
| RFS | MSigDB_TFs | TGGNNNNNNKCCAR_UNKNOWN | 8 | 0.001 | 0.017 |
| RFS | MSigDB_TFs | CAGGTG_E12_Q6 | 54 | 0.003 | 0.043 |
| RFS | MSigDB_TFs | WTGAAAT_UNKNOWN | 14 | 0.006 | 0.046 |
| RFS | MSigDB_TFs | NKX62_Q2 | 6 | 0.008 | 0.088 |
| RFS | MSigDB_TFs | NF1_Q6 | 5 | 0.010 | 0.025 |
| RFS | MSigDB_TFs | CEBPGAMMA_Q6 | 7 | 0.018 | 0.068 |
| RFS | MSigDB_TFs | CTTTAAR_UNKNOWN | 18 | 0.020 | 0.050 |
| RFS | MSigDB_TFs | CAGCTG_AP4_Q5 | 23 | 0.021 | 0.063 |
| RFS | MSigDB_TFs | SRY_02 | 5 | 0.022 | 0.025 |
| RFS | MSigDB_TFs | AP2_Q6 | 5 | 0.026 | 0.037 |
| RFS | MSigDB_TFs | RORA1_01 | 5 | 0.028 | 0.046 |
| RFS | MSigDB_TFs | GATA1_01 | 5 | 0.043 | 0.025 |
| RFS | MSigDB_TFs | AP4_Q5 | 9 | 0.048 | 0.085 |
| RFS | Predicted_TFs | E2F4_MA0470.1 | 6 | 0.048 | 0.067 |
| MetDx | GO | polymeric cytoskeletal fiber | 9 | 0.010 | 0.003 |
| MetDx | GO | ephrin receptor signaling pathway | 5 | 0.013 | 0.071 |
| MetDx | GO | digestive tract development | 8 | 0.014 | 0.086 |
| MetDx | GO | digestive system development | 8 | 0.014 | 0.086 |
| MetDx | GO | system process | 29 | 0.015 | 0.061 |
| MetDx | GO | regulation of body fluid levels | 7 | 0.016 | 0.084 |
| MetDx | GO | supramolecular complex | 11 | 0.019 | 0.006 |
| MetDx | GO | supramolecular polymer | 11 | 0.019 | 0.006 |
| MetDx | GO | supramolecular fiber | 11 | 0.019 | 0.006 |
| MetDx | GO | establishment or maintenance of cell polarity | 6 | 0.027 | 0.021 |
| MetDx | GO | positive regulation of cell adhesion | 6 | 0.033 | 0.042 |
| MetDx | GO | microtubule | 5 | 0.035 | 0.010 |
| MetDx | GO | blood coagulation | 5 | 0.036 | 0.064 |
| MetDx | GO | hemostasis | 5 | 0.036 | 0.064 |
| MetDx | GO | coagulation | 5 | 0.036 | 0.064 |
| MetDx | GO | neuron recognition | 5 | 0.045 | 0.071 |
| MetDx | MSigDB_TFs | RGAANNTTC_HSF1_01 | 10 | 0.025 | 0.004 |
| MetDx | MSigDB_TFs | GATA6_01 | 8 | 0.028 | 0.030 |
| MetDx | MSigDB_TFs | GCTNWTTGK_UNKNOWN | 8 | 0.029 | 0.039 |
| MetDx | MSigDB_TFs | HEB_Q6 | 6 | 0.030 | 0.041 |
| MetDx | MSigDB_TFs | RGAGGAARY_PU1_Q6 | 5 | 0.047 | 0.093 |

**Table S7.** Differential analysis of the global methylation patterns and three outcome signatures in the clinical and *in vitro* datasets. The fraction of differentially methylated ($p < 0.05$) CpG sites between the two primary cluster groups **(Fig. 7)** that were concordantly hypo / hyper methylated between the *in vitro* and each clinical dataset are shown.

| Profile | GDSC v. TARGET (%) | GDSC v. JNCCRI (%) | GDSC v. NY (%) |
|---|---|---|---|
| Global | 99.27 | 86.14 | 63.76 |
| RFS | 95.45 | 100 | 100 |
| CR | 100 | 100 | 100 |
| MetDx | 81.97 | 71.43 | 63.76 |

**Table S8.** Correlations between methylation of the outcome signature CpG sites and *in vitro* aggressiveness metrics. The fraction of probes strongly ($|r| < 0.6$) correlated with the aggressiveness metrics is presented.

| Profile | Tumorigenicity | Colony forming | Invasion | Migration | Proliferation |
|---|---|---|---|---|---|
| RFS | 11.5 | 15.5 | 16.0* | 16.0* | 17.9* |
| CR | 5.1 | 34.8* | 6.1 | 7.5 | 9.1 |
| MetDx | 12.3 | 17.6 | 15.5* | 14.7* | 11.2 |

*Permutation p value ≤ 0.1

**Table S9.** Correlations between methylation of the outcome signature CpG sites and *in vitro* response to standard chemotherapy. The fraction of probes strongly ($|r| < 0.6$) correlated with standard chemotherapeutics and the fraction of strong correlations which are positive are presented. MAP was not tested as a combination treatment, so combined MAP response was assessed by combining methotrexate, doxorubicin, and cisplatin response metrics.

| **Profile** | **Cisplatin** | | **Doxorubicin** | | **Methotrexate** | | **MAP** | |
|---|---|---|---|---|---|---|---|---|
| | **% strong corr.** | **% positive** | **% strong corr.** | **% positive** | **% strong corr.** | **% positive** | **% strong corr.** | **% positive** |
| RFS | 2.7 | 50.0 | 6.7* | 92.0 | 10.9 | 73.2 | 3.7* | 85.7 |
| CR | 1.1 | 50.0 | 5.3 | 100 | 5.3 | 30.0 | 2.1 | 75.0 |

*Permutation p value ≤ 0.1

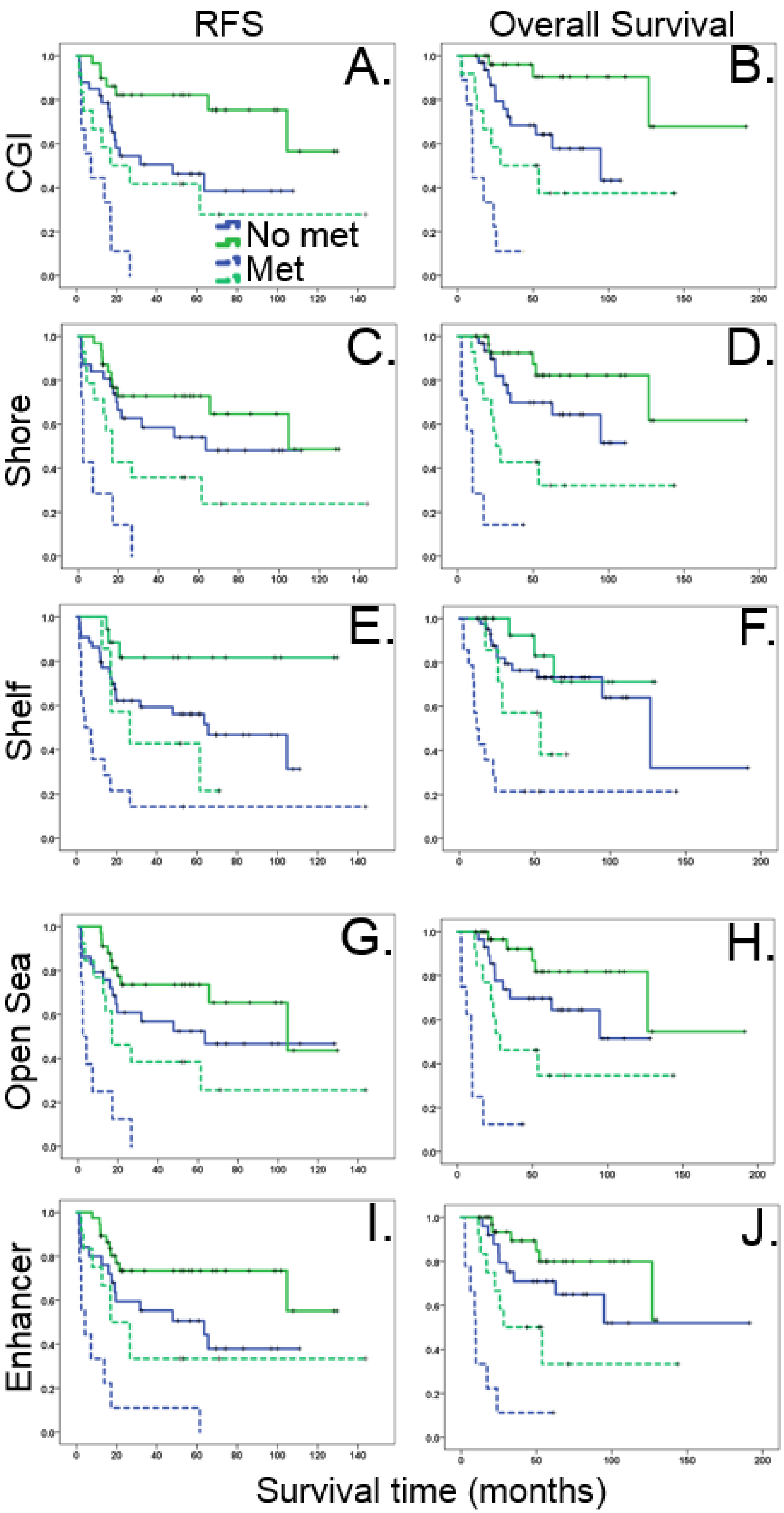


**Figure S1.** Survival analysis of the two main hierarchical clustering groups generated using each of the genomic regions when stratified for metastasis at the time of diagnosis. CGI region: RFS (**A**, pooled p = 0.002) OS (**B**, pooled p = 0.001). Shore region: RFS (**C**, pooled p = 0.036), OS (**D**, pooled p = 0.005). Shelf region: RFS (**E**, pooled p = 0.006), OS (**F**, pooled p = 0.044). Open Sea region: RFS (**G**, pooled p = 0.011), OS (**H**, pooled p = 0.002). Enhancer region: RFS (**I**, pooled p = 0.003), OS (**J**, pooled p = 0.008).

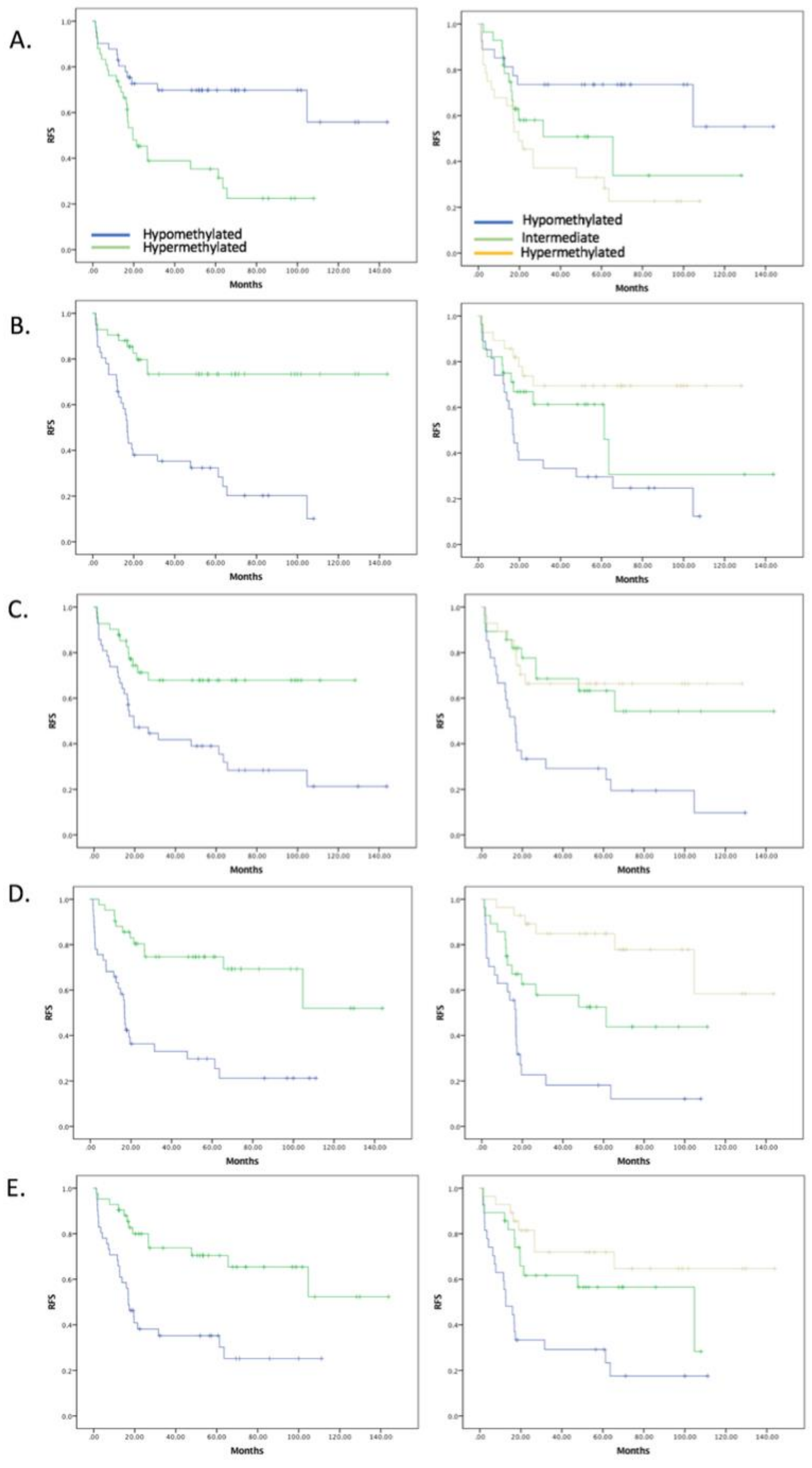


**Figure S2.** RFS analysis of patient risk groups generated with methylation of single CpG site from each analyzed genomic region. Patient risk groups were generated by methylation level (median split and terciles). **A)** CGI region CpG site cg19848683 (MECOM), 2-group log-rank p value = 0.005, 3-group log-rank p value = 0.003. **B)** Shore region, CpG site cg04461028 (HDAC4), 2-group log-rank p value < 0.001, 3-group log-rank p value = 0.004. **C)** Shelf region, CpG site cg06835212 (MEF2C) 2-group log-rank p value = 0.002, 3-group log-rank p value < 0.001. **D)** Open Sea region, CpG site cg15595627 (ANGPT1), 2-group log-rank p value < 0.001, 3-group log-rank p value < 0.001. **E)** Enhancer region, CpG site cg12506775 (TASOR2), 2-group log-rank p value < 0.001, 3-group log-rank p value < 0.001.

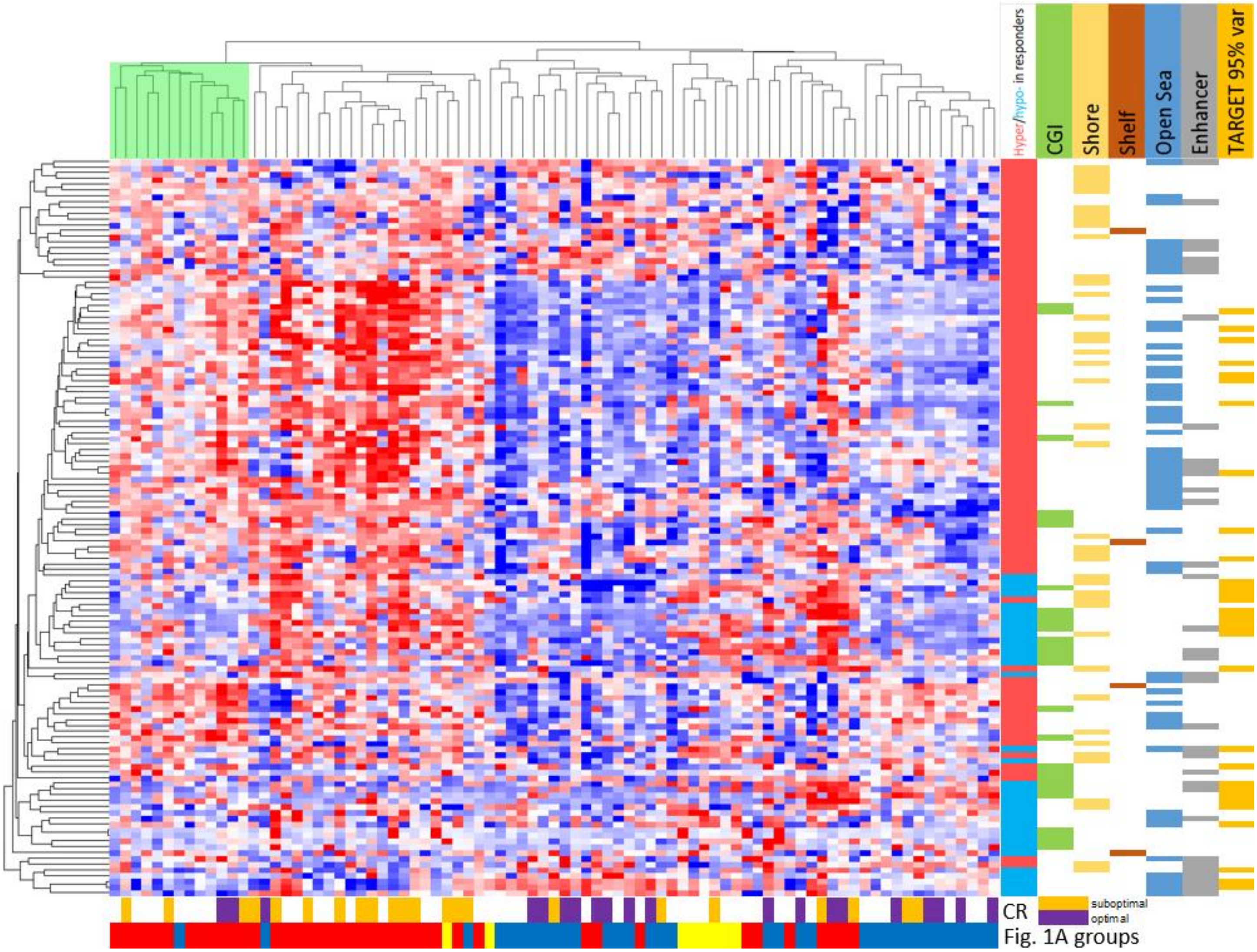


**Figure S3.** Analysis of the EPIMMUNE lung cancer methylation signature. Hierarchical clustering and the heatmap were generated using M values of the 128 EPIMMUNE CpG sites mapped to the TARGET osteosarcoma dataset (two group cluster stability R index = 0.745). Green highlighted samples have a methylation profile predictive of optimal immune checkpoint inhibitor response in the EPIMMUNE discovery cohort. Sites hyper- (red) or hypo- (blue) methylated in immune checkpoint inhibitor responders in the EPIMMUNE study discovery cohort are displayed to the right of the heatmap (leftmost color bar). Genomic regions (middle color bars) and sites passing the 95% variance filtering criteria used for discovery analysis in the TARGET dataset (rightmost color bar) are also displayed. Color bars below the heatmap annotate TARGET osteosarcoma sample response to chemotherapy, and cluster group membership from the genome-wide clustering analysis **(Fig. 1A)**.
